# Single-cell transcriptomic atlas of individuals receiving inactivated COVID-19 vaccines reveals distinct immunological responses between vaccine and natural SARS-CoV-2 infection

**DOI:** 10.1101/2021.08.30.21262863

**Authors:** Yi Wang, Xiaoxia Wang, Laurence Don Wai Luu, Jieqiong Li, Xiaodai Cui, Hailan Yao, Xin Zhang, Shaojin Chen, Jin Fu, Licheng Wang, Chongzhen Wang, Rui Yuan, Qingguo Cai, Xiaolan Huang, Junfei Huang, Wenjian Xu, Shijun Li, Xiong Zhu, Jun Tai

## Abstract

To control the ongoing COVID-19 pandemic, CoronaVac (Sinovac), an inactivated vaccine, has been granted emergency use authorization by many countries. However, the underlying mechanisms of the inactivated COVID-19 vaccine-induced immune response remain unclear, and little is known about its features compared to SARS-CoV-2 infection. Here, we implemented single-cell RNA sequencing (scRNA-seq) to profile longitudinally collected PBMCs (peripheral blood mononuclear cells) in six individuals immunized with CoronaVac and compared these to the profiles of COVID-19 infected patients from a Single Cell Consortium. Both inactivated vaccines and SARS-CoV-2 infection drove changes in immune cell type proportions, caused B cell activation and differentiation, and induced the expression of genes associated with antibody production in the plasma. The inactivated vaccine and SARS-COV-2 infection also caused alterations in peripheral immune activity such as interferon response, inflammatory cytokine expression, innate immune cell apoptosis and migration, effector T cell exhaustion and cytotoxicity, however, the magnitude of change was greater in COVID-19 patients, especially those with severe disease, than in immunized individuals. Further analyses revealed a distinct peripheral immune cell phenotype associated with CoronaVac immunization (HLA class II upregulation and *IL21R* upregulation in naïve B cells) versus SARS-CoV-2 infection (HLA class II downregulation and *IL21R* downregulation in naïve B cells severe disease). There were also differences in the expression of important genes associated with proinflammatory cytokines and thrombosis. In conclusion, this study provides a single-cell atlas of the systemic immune response to CoronaVac immunization and reveals distinct immune responses between inactivated vaccines and SARS-CoV-2 infection.

## Introduction

COVID-19 (coronavirus disease 2019), caused by SARS-CoV-2 (severe acute respiratory syndrome coronavirus 2), is an unprecedented threat to global public health and has rapidly spread throughout the world (Cevik et al., 2020). COVID-19 has led to high mortality and morbidity worldwide, and as of Aug 24, 2021, 213,752,662 laboratory-confirmed cases of SARS-CoV-2 infection have been reported, resulting in 4,459,381 deaths (WHO COVID-19 Dashboard). As there are no effective drugs available at this time against COVID-19, safe and effective COVID-19 vaccines are urgently required to control the pandemic and reduce the global burden of SARS-CoV-2 (Mehra et al., 2020).

Various candidate vaccines including inactivated viral vaccines, live attenuated vaccines, nucleic acid vaccines, viral-vectored vaccines, and protein or peptide subunit vaccines are being rapidly developed, tested, and granted approval for emergency use (Amanat and Krammer, 2020). Each vaccine has advantages and disadvantages and these have been reviewed elsewhere (Dong et al., 2020; Poland et al., 2020). Among these candidate vaccines, the inactivated COVID-19 vaccines are among one of the most widely used and well developed vaccines due to their ease of production and scale-up, and relatively low cost. They are produced by growing SARS-CoV-2 in cell culture (e.g., Vero cells), followed by chemical inactivation of the virus (Krammer, 2020). Inactivated vaccines present the whole SARS-COV-2 virus for immune recognition, thus the immune responses are likely to target not only the unique protein (e.g., S protein) of the virus but also matrix, nucleoprotein and envelope (Krammer, 2020). Moreover, the inactivated vaccines exhibit stable expression of conformation-dependent antigenic epitopes, and also offer advantages in a variety of different populations (e.g., those with degrees of immune senescence) (Iversen and Bavari, 2021b).

CoronaVac (initially known as PiCoVacc) from Sinovac, is a leading Chinese COVID-19 vaccine and was devised with β-propiolactone as an inactivating agent and formulated with aluminum hydroxide as an adjuvant (Gao et al., 2020). The inactivated CoronaVac vaccine is a whole-virus preparation administered in a two-dose regimen (at day 0 and day>21), and the immunogenicity, safety and tolerability have been assessed in different populations, including children and adolescent aged 3-17 years old (Han et al., 2021), adults aged 18-59 (Zhang et al., 2021), and adults aged 60 years and older (Wu et al., 2021). Within the scope of combating the SARS-CoV-2 pandemic, CoronaVac has been granted an emergency use authorization by Chinese authorities in July, 2020 (Poland et al., 2020), and a host of others countries such as Turkey, Chile, Brazil, Indonesia, etc. (Bayram et al.; Fortner and Schumacher, 2021; Muena et al., 2021; Shervani et al., 2020).

Currently, the knowledge about the immunity generated by COVID-19 vaccines (including the inactivated vaccines) are limited with researchers understanding less about this than about immunity to natural SARS-CoV-2 infection. Although clinical trial data have demonstrated that the current COVID-19 vaccines approved (including CoronaVac) can elicit immunity with a high degree of safety, efficacy and tolerability, much remains to be learned concerning the genetic drivers of COVID-19 vaccine-elicited humoral and/or cellular immunity, defining detailed targets of the immune response at the epitope level, and characterizing the B-cell receptor and T-cell receptor repertoire induced by COVID-19 vaccines (Poland et al., 2020). Inactivated vaccines (such as CoronaVac) have been shown to keep the immunogenicity of the SARS-CoV-2 virus, and can elicit an immune response, however whether there is a distinct immune response landscape between natural SARS-CoV-2 infection and the inactivated COVID-19 vaccine remains unclear.

Here, we implemented scRNA-seq (single-cell RNA sequencing) to obtain a comprehensive and unbiased visualization of PBMCs (Peripheral blood mononuclear cells) from healthy adults immunized with the inactivated COVID-19 vaccine, CoronaVac, at 3 pivotal time points, including day 0 (before vaccination), day 21 after the first dose and day 14 after the second dose (**Figure 1A**). This study provides a high-resolution transcriptomic landscape of PBMCs during the immune response to CoronaVac immunization, which will foster a better understanding of the protective immune response generated by inactivated COVID-19 vaccines. Then, we compared this data to the reported profiles from a Single Cell Consortium for COVID-19 (Ren et al., 2021), revealing the distinct immune features between natural SARS-CoV-2 infection and the inactivated COVID-19 vaccine.

**Fig 1.**
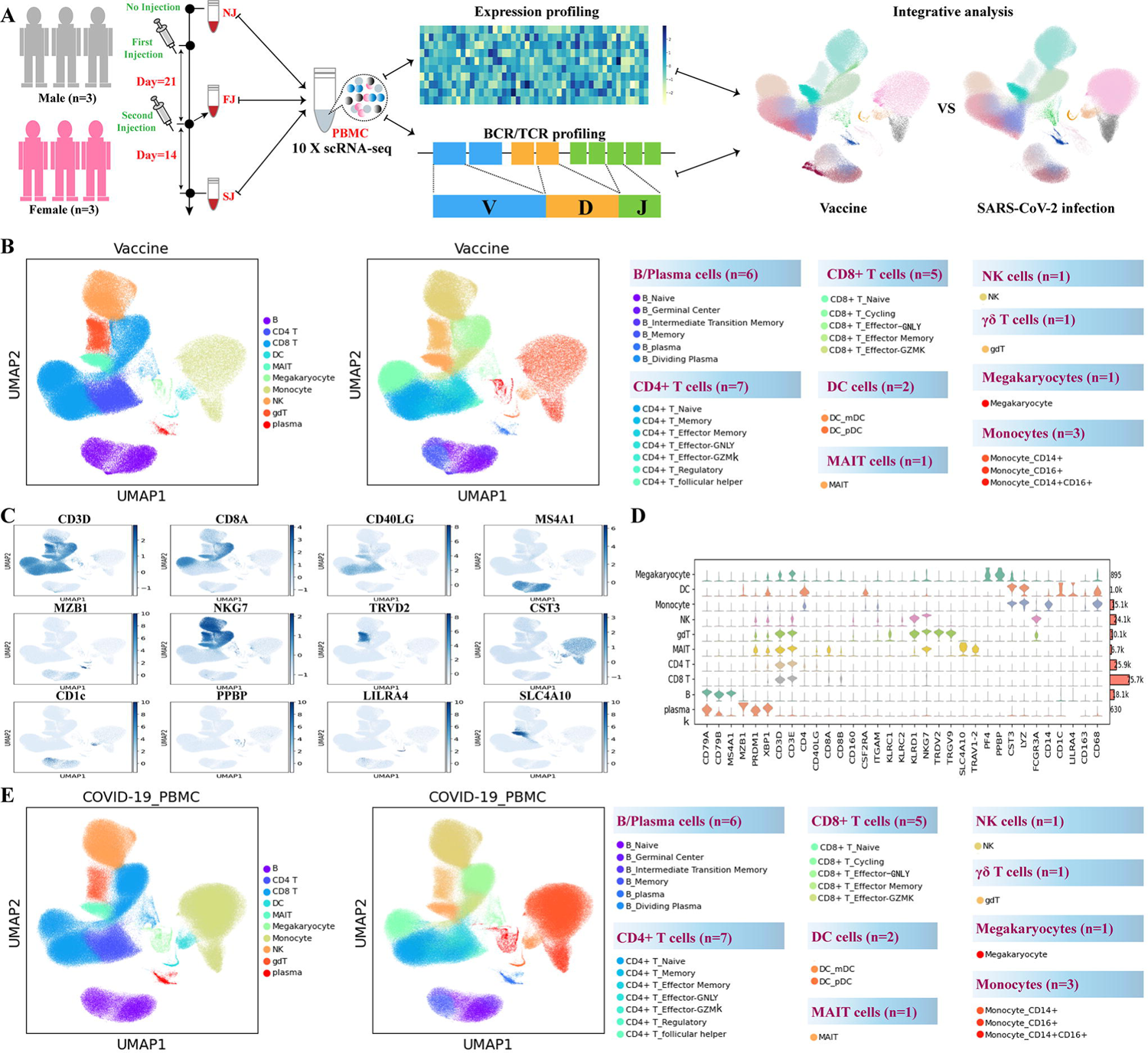
Study design and overall results of single-cell transcriptomic profiling of PBMCs isolated from vaccine recipients without COVID-19 infection. **A**, A schematic diagram of the overall study design. The PBMCs from six recipients, 3 male and 3 female adults, and across three conditions were subjected to scRNA-seq gene expression profiling, TCR and BCR profiling analyses. The data set was integrated with a published COVID-19 scRNA-seq data set comprised of 64 fresh PBMC samples. **B**. Cell populations identified and 2-D visualization. The UMAP projection of 180k single cell transcriptomes from NJ (n = 6), FJ (n = 6) and SJ (n = 6) samples, showing the presence of 10 major clusters and 27 smaller clusters with their respective colors. Each dot corresponds to a single cell, colored according to the annotated major cell type ( left panel) or subcelltype ( right panel). **C**. Canonical single cell RNA markers were used to label major clusters by cell identity as expression level on the UMAP plot. Cells are colored according to log transformed and normalized expression levels of twelve genes (CD3D, CD8A, CD40LG, etc.). **D.** Expression distribution of cell identity specific RNA markers of vaccine cohort samples. The rows represent 10 cell clusters labeled with different colors and the columns represent log transformed gene expression of the RNAs. The distribution of a gene in a cluster is shown as one small violin plot. **E.** Similar to Fig 1B, Cell populations identified and 2-D visualization of 410k single cell transcriptomes from Cont (Control), Conv (Convalescence), Mild (Mild), and Seve (Severe) samples from Ren et al..

## Results

### Single-cell transcription profiling of PBMCs

To identify features of the immunological hallmarks from individuals receiving inactivated COVID-19 vaccines (CoronaVac), the droplet-based scRNA-seq (10 X Genomics) was conducted for studying the transcriptomic profiles of PBMCs, which were longitudinally collected from six individuals at three pivotal time points (**Fig 1A**). Single-cell B cell receptor (BCR) and T cell receptor (TCR) sequencing were also performed for each sample. According to the time points, these samples were classified into three conditions: no injection (NJ, day=0, PMBCs were collected before vaccination), first injection (FJ, Day=21, PBMCs were collected at day 21 after the first dose) and second injection (SJ, Day=35 PBMCs were collected at day 14 after second dose). The associated metadata of the six individuals enrolled and the three conditions are detailed in **Supplementary Table S1**. After the single-cell analysis pipeline (Refer to Methods), we obtained ∼0.895 billion unique transcripts belonging to 178268 cells from the PBMCs of vaccinated individuals. Among these cells, 60783 cells (34.1%) were from the NJ conditions, 47451 cells (26.6%) were from the FJ condition, and 70034 cells (39.3%) from the SJ condition. Next, we integrated all high-quality cells into an unbatched and comparable dataset, which was subjected to principal component analysis after correction for read depth and mitochondrial read counts (**Fig S1A-B**)

To reveal immune cell populations in individuals administered with inactivated COVID-19 vaccines, the graph-based clustering of UMAP (uniform manifold approximation and projection) was performed. According to the expression of canonical cell-type markers, we identified 10 major cell types (**Fig 1B (left), C-D**; **S1A-B**): B cells (CD79A^+^CD79B^+^MS4A^+^), plasma cells (XBP1^+^MZB1^+^), γδ T cells (TRDV1^+^TRDV9^+^), natural killer (NK) cells (CD11b^+^NKG7^+^KLRD1^+^NKG2A^+^), CD4^+^ T cells (CD3D^+^CD3E^+^CD40LG^+^), CD8^+^ T cells (CD3D^+^CD3E^+^CD8A^+^CD8B^+^), mucosal-associated invariant T (MAIT) cells (CD3D^+^CD3E^+^SLC4A10^+^), monocytes (CST3^+^LYZ^+^CD68+), dendritic cells (CST3^+^LYZ^+^CD163^+^) and megakaryocytes (CST3^+^LYZ^+^PPBP^+^). At the more granular level, we identified 27 different cell subtypes (**Fig 1B (right)**, **S1A-B**). Likewise, we also successfully identified 10 major cell types (**Fig 1E** (**Left**)) and 27 cell subtypes (**Fig 1E** (**Right**)) for the PBMCs samples reported by a Single Cell Consortium for COVID-19 (Ren et al., 2021) (**Fig S1C-F**). As such, the composition of cell subpopulations in peripheral blood from individuals with COVID-19 vaccine and COVID-19 patients were clearly defined (**Fig 1B**, **E**; **S1G**).

### Differences in major cell type compositions across conditions

We firstly uncovered the differences in cell composition (10 major cell types) across two conditions (FJ and SJ) and then compared that with NJ. According to scRNA-seq data (**Fig 2A-D**), we calculated the relative percentage of 10 major cell types in the PBMCs of each participant at 3 conditions. The relative abundance of CD4+ T cells, CD8+ T cells, γδ T cells, NK cells and MAIT cells at FJ and SJ conditions remained similar when compared with the NJ condition (**Fig 2D**). The relative percentage of B cells appeared to increase in FJ and SJ conditions in comparison with NJ condition, implying that the change in B cells may be related with the humoral immune response after vaccination (**Fig 2D**). The proportions of dendritic cells (DCs), monocytes (Mono) and megakaryocytes (Mega) increased after vaccination (**Fig 2D**). Increased DCs may be involved in antigen presentation to stimulate the immune response to CoronaVac, while increased Mono and Mega may be involved with the potential inflammatory response after vaccination. Of note, the percentage of plasma cells did not significantly increase in the FJ and SJ conditions (**Fig 2D**). This may be due to plasma cells requiring a strong level of continuous antigen stimulation, or that the plasma cell level had decreased or was restored when the PBMC samples were collected at the FJ (at day 21 after first dose) and SJ (at day 14 after first dose) conditions.

**Fig 2.**
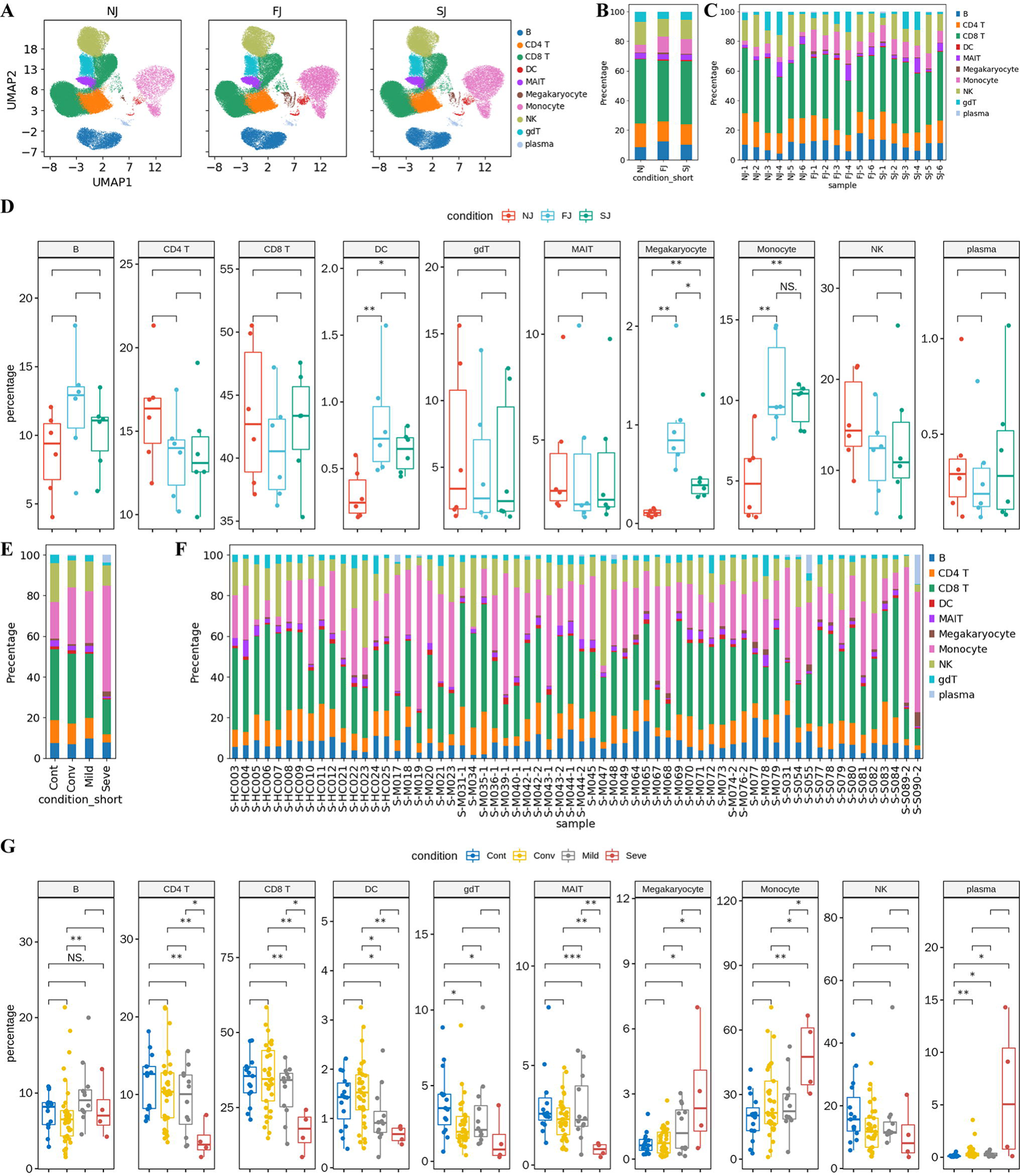
Comparison of cell composition across sample groups. **A.** UMAP projection of the single cells from NJ, FJ and SJ conditions. Each dot corresponds to a single cell, colored by its major cell type. **B.** Average proportion of each major cell type derived from NJ, FJ and SJ groups. **C.** Proportion of each major cell type derived from each NJ, FJ and SJ individual sample. **D.** The box plot shows cell compositions of NJ, FJ and SJ conditions at a single sample level. Condition preference of each cluster compared side by side. y axis, percentage of cell types. Boxes are colored by conditions. Each plot panel represents one cell type. **E.** Average proportion of each major cell type derived from Cont, Conv, Mild, and Seve conditions from the COVID-19 infection cohort. **F.** Proportion of each major cell type derived from individual samples of Cont, Conv, Mild, and Seve groups from the COVID-19 infection cohort **G.** The box plot shows cell compositions of Cont, Conv, Mild, and Seve conditions from the COVID-19 infection cohort at a single sample level. In 2D and 2G, all pairwise differences with P < 0.05 using two-sided unpaired Mann–Whitney U-test are marked to show significance levels.

Next, we compared the cell composition between vaccine and natural SARS-CoV-2 infection. Patients with COVID-19 (n=64) were classified into four conditions: control (Cont; n=15), mild (mild; n=12), severe (seve; n=4) and convalescent (conv; n=33) (**Fig 2E-G, Fig S2A**). After SARS-CoV-2 infection, the proportion of innate immune cells, including NK cells, γδ T cells, MAIT cells and DCs, decreased with disease severity (**Fig 2E-G**). This trend was different to what was observed with vaccines (**Fig 2D, G; S2B-D**). Similar with vaccination, the relative abundance of monocytes and megakaryocytes in COVID-19 patients increased with disease severity, and the relative percentage of these cells later declined in Conv conditions (**Fig 2G**). Unlike the inactivated COVID-19 vaccine, a decrease in CD4^+^ T and CD8^+^ T cells were observed in COVID-19 infected patients, and this was related with disease severity (**Fig 2D, G; S2D**). A slight increase in B and plasma cell levels were observed after SARS-CoV-2 mild infection, whereas a massive increase in plasma cells was observed in the Seve condition (**Fig G**; **S2D**). These data suggested that both vaccination with CoranaVac and natural SARS-CoV-2 infection can cause changes in the cell compositions of PBMC.

### Features of B cell subsets across samples

To reveal the dynamic changes in different B cell subtypes in immunized (**Figure 3A-E, Fig S3A**) and infected individuals (**Fig 3F-G**, **S3B-E**), we classified B cells into 6 subsets according to the distribution and expression of classical subtype markers. We successfully identified one naïve B subcluster (MS4A1^+^IGHD^+^), one memory B subcluster (MS4A1^+^CD27^+^), one germinal center B subcluster (MS4A1^+^NEIL1^+^), one intermediated transition memory B subcluster (Intermediate memory B, MS4A1^+^IGHD^+^CD27^+^), one plasma B subcluster (MZB^+^CD38^+^) and one proliferating plasma B subsets (MZB^+^CD38^+^MKI67+).

**Fig 3.**
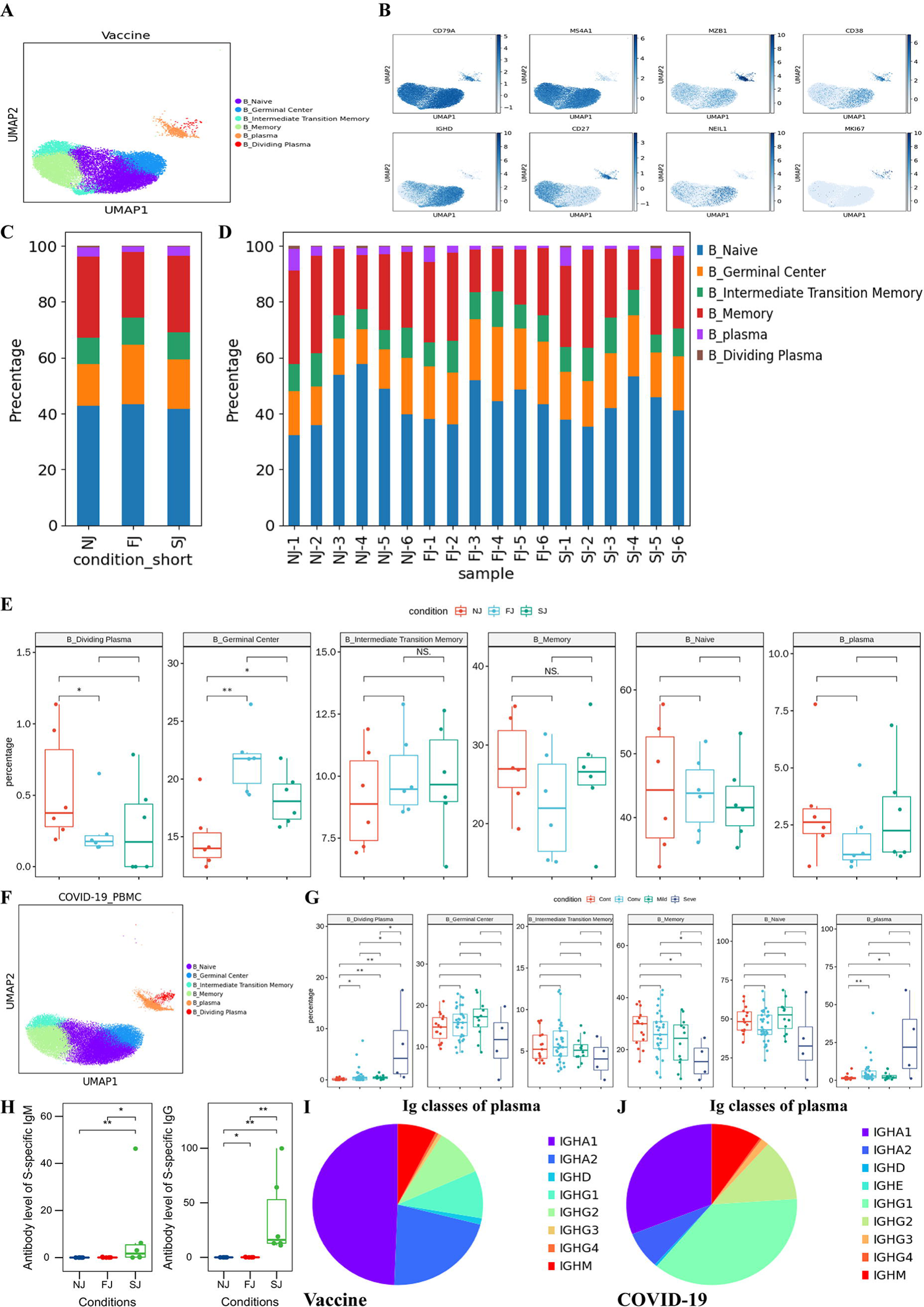
Characterization of B cell composition differences in individuals across vaccination and infection conditions. **A.** UMAP projection of all B cells from NJ, FJ and SJ conditions. Each dot corresponds to a single cell, colored by its cell subtype. **B.** Expression levels of canonical B cell RNA markers were used to identify and label major cell clusters on the UMAP plot. Cells are colored according to log transformed and normalized expression levels of eight genes. Cells are from NJ, FJ and SJ conditions **C.** Average proportion of each B cell subtype derived from NJ, FJ and SJ groups. **D.** Proportion of each B cell subtype derived from NJ, FJ and SJ individual samples. **E.** The box plot shows the composition of B cells before (NJ) and after vaccination (FJ, SJ) at a single sample level. **F.** UMAP projection of all B cells from Cont, Conv, Mild and Seve conditions. Each dot corresponds to a single cell, colored by its cell subtype. **G.** Proportion of each B cell subtype derived from Cont, Conv, Mild and Seve individual samples. **H.** Antibody levels of IgM and IgG at NJ, FJ and SJ conditions in serum. **I.** The composition of Ig classes in the vaccine cohort identified by BCR single cell sequencing. **J.** The composition of Ig classes in the COVID-19 infected cohort identified by BCR single cell sequencing. All pairwise differences with P < 0.05 using two-sided unpaired Mann–Whitney U-test are marked to show significance levels.

The general patterns of B/plasma cells were compared across conditions. The relative percentages of an active state B subtype (germinal center B) significantly increased after vaccination, suggesting that B cells may be activated after vaccination (**Fig 3E**), Other B cell subsets, including naïve B cells, memory B cells and intermediate memory B cells, remained similar across the three vaccination conditions (NJ, FJ and SJ) (**Fig 3E**), suggesting that the inactivated COVID-19 vaccine has a relatively low impact on the composition of these B cell subsets. Likewise, SARS-CoV-2 infection also had relatively low impact on the composition of these B cell subsets, with only the memory B subtype decreased in severe COVID-19 patients (**Fig 3G**). Increased plasma and dividing plasma cells were observed in COVID-19 patients with the percentage of plasma cells in severe COVID-19 patients reaching 15% while mild COVID-19 patients only reached 3% (**Fig 3G**) (Ren et al., 2021), suggesting that antibody production may be stronger in severe COVID-19 patients, reminiscent of previous findings that higher antibody titers are related with worse clinical outcomes (Long et al., 2020; Okba et al., 2020). Although the levels of plasma cells and dividing plasma cells did not significantly increase after vaccination, the levels of neutralizing antibodies (anti-S-RDB-specific antibody) did significantly increase after the second injection for all 6 individuals (**Fig 3H**, **Fig S3G**, **Table S2**). Interestingly, the levels of neutralizing antibody did not significantly increase after the first dose of CoronaVac (**Fig 3H**, **Fig S3G**, **Table S2**) and significant production of neutralizing antibody was only obtained after the second dose. This implies that two doses of CoronaVac are required for efficient seroconversion.

After vaccination, the plasma cells in PBMCs had highly expressed genes which encode the constant regions of immunoglobulin G1 (Ig G1), IgG2, IgA1 or IgA2. This correlates with their function in secreting antigen-specific antibodies and implies that the serum of immunized individuals may have had high titers of SARS-CoV-2-specific antibodies (**Fig 3H-I, S3G**). These findings were also observed in COVID-19 patients (**Fig 3J**), and is consistent with previous studies showing high titers of antigen-specific antibodies in the serum of COVID-19 patients (Ni et al., 2020; Ren et al., 2021). Similar to natural SARS-CoV-2 infection, genes encoding IgA1 and IgA2 were activated by CoronaVac, and this finding has also been observed in influenza vaccines (Neu et al., 2019).

### Transcriptomic changes in B cells after vaccination and SARS-CoV-2 infection

To investigate differential transcriptomic changes in B/plasma cells after vaccination, we compared the expression profiles of B/plasma cells in FJ or SJ conditions with the NJ condition. As expected, genes involved in B cell activation, adaptive immune response, response to interferon, and antigen processing and presentation were specifically enriched in B cells after vaccination (**Fig 4A**). This suggests that the B cells were responding to the inactivated COVID-19 vaccine. For SARS-CoV-2 infection, genes involved in defense response to virus and interferon signaling pathways were the most highly upregulated in the B cell subset (**Fig S4A**). Notably, we observed that genes associated with the “IFN response” were enriched in both post-vaccination samples and COVID-19 patients (**Fig 4A** and **Fig S4A**).

**Fig 4.**
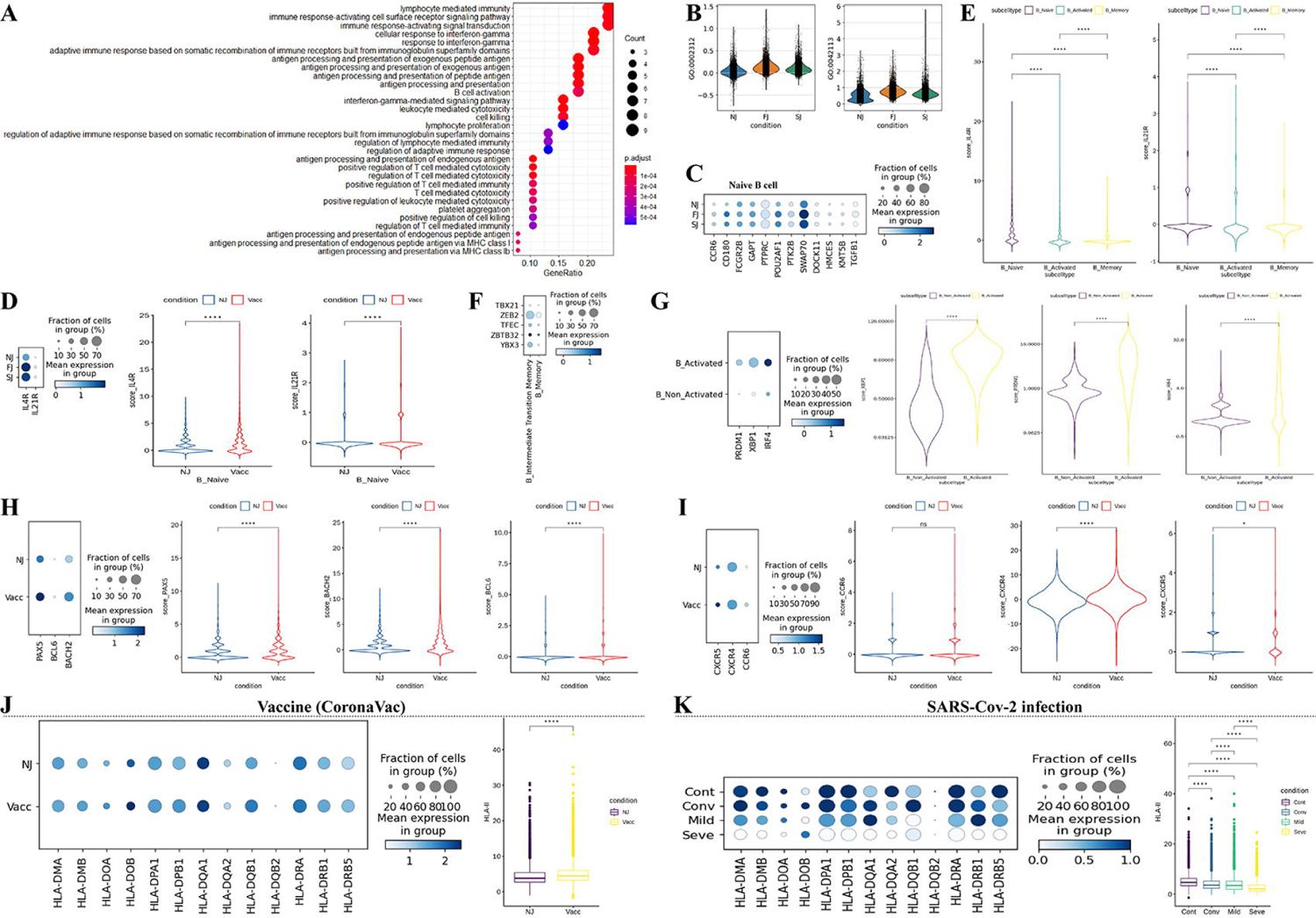
Characterization of gene expression differences in B cells in individuals across vaccination and infection conditions. **A.** GO enrichment analysis of DEGs identified by comparing before- and after-vaccination conditions. DEGs refer to genes with Benjamini–Hochberg adjusted P value (two-sided unpaired Mann–Whitney U-test) ≤0.01 and average log2 fold change ≥1 in both FJ/NJ and SJ/NJ comparisons. **B.** Violin plots of B cell expression activities in three vaccine conditions. Cells are grouped and colored by conditions. Y axis represents the normalized expression score of gene sets related to B_CELL_ACTIVATION_INVOLVED_IN_IMMUNE_RESPONSE (GO:0002312) and B_CELL_ACTIVATION (GO:0042113) **C.** Dot plots of the gene expression level of naïve B cells in three vaccine conditions. Rows represent conditions; columns represent genes. Dots are colored by mean expression levels in each condition. **D.** (Left) Dot plots of *IL4R* and *IL21R* expression level of naïve B cells between pre-vaccination and post-vaccination. Rows represent conditions; columns represent genes. Dots are colored by mean expression levels in each condition. (Right) Violin plots show the expression level of the two genes in before- and after-vaccination conditions. **E.** Violin plots of the expression level of *IL4R* and *IL21R* genes in naïve B cells, activated B cells and memory B cells. **F.** Dot plots of gene expression level of memory B and intermediate transition memory B cells in vaccine cohort. Rows represent genes (*TBX21*, *ZEB2*, *TFEC*, *ZBTB32*, *YBX3*); columns represent B cell subtypes. Dots are colored by mean expression levels in each group. ***G.*** *PRDM1*, *XBP1*, *IRF4* gene expression level of activated B cells and other B cells in the vaccine cohort. Dot plots (Left) and violin plots (Right) are used for visualization. ***H.*** *PAX5*, *BCL6* and *BACH2* gene expression level of B cells in before- and after-vaccination conditions. Dot plots (Left) and violin plots (Right) are used for visualization. ***I.*** *CXCR5*, *CXCR4* and *CCR6* gene expression level of B cells in before- and after-vaccination conditions. Dot plots (Left) and violin plots (Right) are used for visualization. **J.** Gene expression level of HLA-II genes in B cells in before- and after-vaccination conditions. Dot plots (Left) show individual genes and violin plots (Right) show normalized average expression of the HLA-II gene set. **K.** Gene expression level of HLA-II genes in B cells in Cont, Conv, Mild and Seve conditions. Dot plots (Left) show individual genes and violin plots (Right) show normalized average expression of the HLA-II gene set. All pairwise differences with P < 0.05 using two-sided unpaired Mann–Whitney U-test are marked to show significance levels.

Next, we further examined the expression of important genes (e.g., *PRDM1*, T-bet) that are involved in B/plasma-cell-activation-related processes after vaccination and SARS-CoV-2 infection. Two GO pathways (GO:0002312 and GO:0042113) that were related to the activation of B cells were significantly enriched after vaccination (**Fig 4B**). Several genes (e.g., *PTPRC*, *HMCES* and *SWAP70*) involved in the B cell activation GO pathways (GO:0002312 and GO:0042113), were highly upregulated in the naïve B cell subtype, implying activation of naïve B cells after vaccination (**Fig 4C**). In contrast, these three genes (*PTPRC*, *HMCES* and *SWAP70*) were down-regulated in severe COVID-19 patients, suggesting that activation of naïve B cells in these individuals may be impaired (**Fig S4B**). Naïve B cells in vaccinated samples also highly expressed *IL4R* and *IL21R* (**Fig 4D**), but these genes were downregulated in activated and memory B cells (**Fig 4E**). This indicates that naïve B subsets were more responsive than activated and memory B cells to IL-4 (interleukin 4) and IL-21, which regulate class switching to IgG, including IgG1, IgG3 or IgG4 (Horns et al., 2020; Pène et al., 2004). Similar findings for *IL4R* and *IL21R* expression were also observed in SARS-CoV-2 mild infection, while *IL4R* and *IL21R* were significantly downregulated in severe COVID-19 patients (**Fig S4C-D**).

Six transcription factors (TBX21, ZEB2, TFEC, ZBTB32 and YBX3) associated with the activation of memory B cells were highly expressed in intermediate transition memory B cell (also referred to as activated memory B cells) compared to memory B cells (**Fig 4F** and **Fig S4E**) (Horns et al., 2020). TBX21 (also known as T-bet) has been hypothesized to play a key regulatory role in activation and is required for class switching to IgG2 (Wang et al., 2012). This transcription factor showed higher expression in activated memory B cells than memory B cells (**Fig 4F** and **Fig S4E**). A triad of transcription factors, including PRDM1, XBP1 and IRF4, also had increased expression in activated B cells (which encompasses: germinal center B cell, intermediate transition memory B cell, dividing plasma and plasma) from COVID-19 patients and immunized individuals than non-activated B cell (including naïve B cell and memory B cell) (**Fig 4G** and **Fig S4F**). These transcription factors are associated with B-cell-differentiation-related pathways and are required for activating the ASC (antibody-secreting cell) program (Shi et al., 2015). PRDM1 plays a core role in determining and shaping the secretory arm of B cell differentiation and in promoting Ig synthesis. XBP1 is a positively acting transcription factor of the CREB-ATF family that is highly express in plasma cells and is important for increasing protein synthesis in plasma cells (Shaffer et al., 2004). IRF4 is crucial for regulating Ig class-switch recombination, and a previous study has found that sustained and increased concentration of this transcription factor promotes the generation of plasma cells (Ochiai et al., 2013). Finally, three B cell-promoting transcription factors: BACH2, BCL6 and PAX5, showed increased expression in B cells after vaccination (**Fig 4H**) while in severe COVID-19 patients (**Fig S4G**), these transcription factors decreased. BACH2, BCL6 and PAX5 play a key role in determining the fate of B cells during differentiation (Shi et al., 2015).

Interestingly in COVID-19 infected samples, expression of important chemokine receptors such as CXCR5 were significantly decreased, especially for severe samples (**Fig S4H**). However, this loss of chemokine receptors was not observed in immunized samples (**Fig 4I**). Decreased chemokine receptors can impair germinal center reactions and ultimately cause dysregulated humoral immunity responses (Mathew et al., 2020; Okada et al., 2002; Reimer et al., 2017). Our data also observed significant upregulation of HLA class II genes after immunization (**Fig 4J**), implying that there is an enhancement of immune cell crosstalk between the adaptive immune cell classes. However in COVID-19 patients, several HLA class II genes were significantly downregulated, especially for severe COVID-19 patients (**Fig S4I**). This suggests a dysregulation of immune cell crosstalk between the adaptive immune cell classes during infection. Together, these data define the transcriptional hallmarks of CoronaVac-induced B cell activation and clonal expansion and revealed underlying differences in the B cell transcriptome between vaccine-induced immunity and SARS-CoV-2 infection.

### V(D)J gene usage and clonal expansion in B cells after vaccination and SARS-CoV-2 infection

BCR information was detected in all B/plasma subsets and in >80% of cells while clonal expansion (clonal size >10) was observed in memory B cells, intermediate transition memory B cells, dividing plasma cells, and plasma cells (**Fig 5A**, **Fig S5A-B**). The largest proportion of BCR in B cells was the IGHM subtype and the largest proportion of BCR in plasma cells were IGHA1 and IGHG1 (**Fig 5B**). After vaccination, the percentage of IGHM significantly increased in B cells (**Fig 5C, Fig S5A**) while the percentage of IGHA1 decreased in plasma cells (**Fig 5C, S5C, S5D**). The light chain type, IGK and IGL, did not change in B and plasma cells (**Fig 5D**).

**Fig 5.**
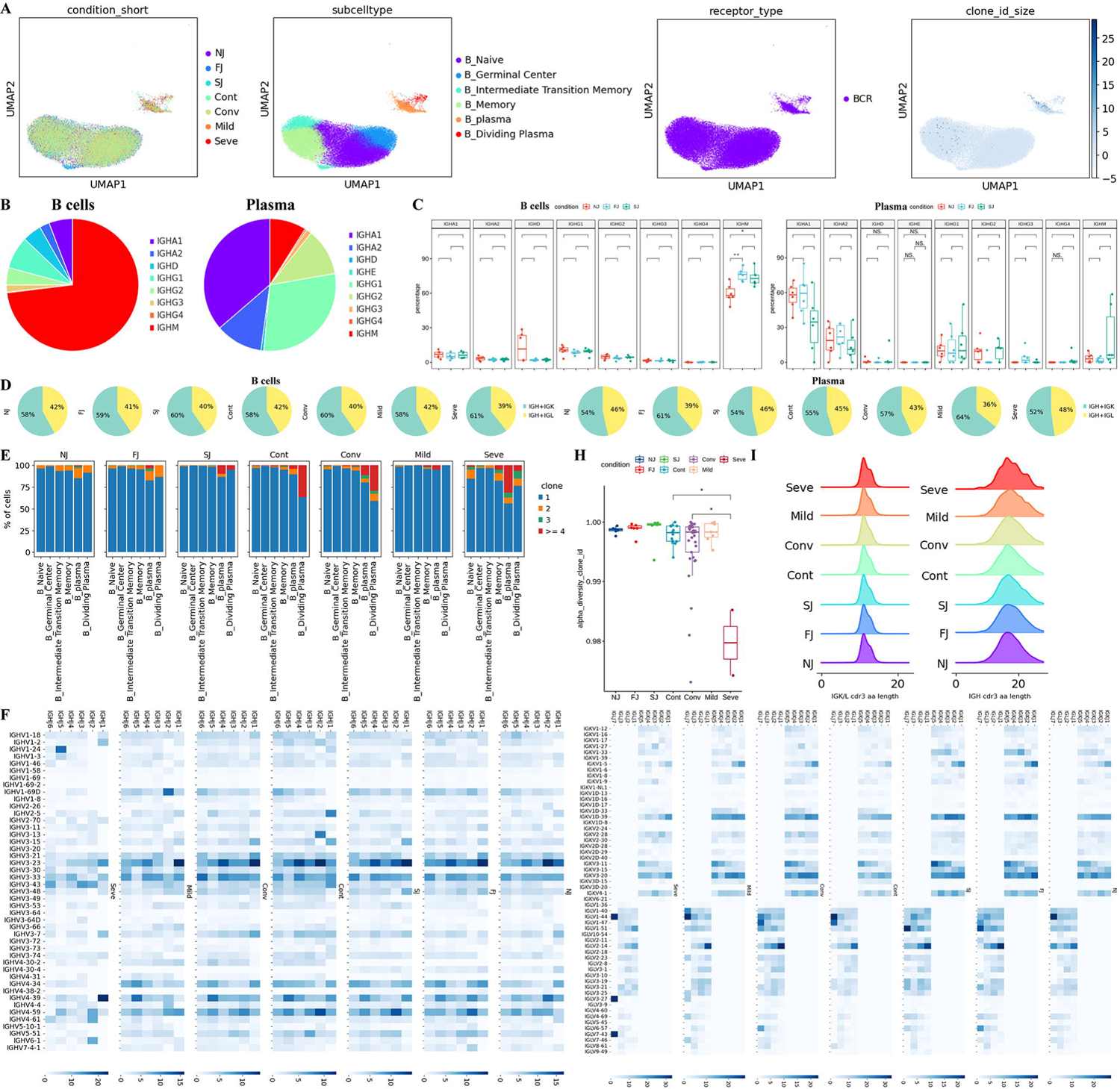
Changes in BCR clones and selective usage of V(D)J genes. **A.** UMAP projection of B cells derived from PBMCs. Cells are colored by conditions (Panel 1), B cell subtypes (Panel 2), if BCR detection was successful (Panel 3) and clone-type expansion size (panel 4) **B.** Pie graph showing the distribution of IGHA, IGHD, IGHG and IGHM in B cells and plasma cells. **C.** Box plot showing the percentages IGHA, IGHD, IGHG and IGHM in B cells and plasma cells under each condition. **D.** Pie graph showing the distribution of light chain IGK and IGL in B cells and plasma cells under each condition. **E.** Stacked bar plots showing the clone state of each B cell subtype in each condition. **F.** Heat maps showing differential IGH/K/L rearrangement. Prevalent IGHV-IGHJ combination pairs (left) and IGKV-IGKJ combination pairs (right) are compared across conditions. Usage percentage are sum normalized by column. **H.** Box plot showing the alpha diversity of clonotypes in each PBMC sample. Data points are colored by condition. **I.** Density curve plots showing the distribution shift of IGK/L and IGH chain CDR3 region length in BCR clone types for each condition.

We compared the clonal expansion of B cell subtypes under different conditions in the vaccine cohort (**Fig 5E**). BCR clonal expansion of plasma cells increased from NJ to FJ to SJ, suggesting that two-doses of CoronaVac induced plasma proliferation of specific BCR clonotypes. In COVID-19 patients, increased clonal expansion in CD8+ T was also observed in severe COVID-19 patients (**Fig 5E**), in agreement with Zhang et al. (Zhang et al., 2020).

BCR diversity, as measured by alpha diversity, showed no change after vaccination but was significantly decreased in severe COVID-19 infections (**Fig 5H**). The length distribution of the CDR3 region was similar for all conditions except COVID-19 severe condition (**Fig5I**). These results suggest that vaccine induced immune protection in very gentle way.

The usage of IGH V(D)J genes across vaccination and infection conditions were compared (**Fig 5F**). The combinations of the 6 IGHJ and >40 IGHV genes demonstrated that CoronaVac induced many changes in the IGH V(D)J genes (**Fig 5F** left panel). The V(D)J pair pattern were significantly altered after two doses of CoronaVac. For example, the most prevalent pair in NJ was IGHJ2/IGHV3-23 which shifted to IGHJ2/IGHV4-59 and IGHJ1/IGHV3-23 following vaccination. The percentage of IGHJ1/IGHV3-43 and IGHJ1/IGHV3-15 also increased after vaccination. In addition, we also analyzed the usage of IGK/L V(D)J genes (**Fig 5F** right panel) and observed that the V(D)J pair pattern were also altered after two doses of CoronaVac. For example, IGKJ5/IGKV3-11, IGLJ1/IGKV2-14 and IGLJ7/IGKV1-51 were all increased after vaccination. Interestingly, IGLJ1/IGKV2-14 and IGLJ7/IGKV1-51 were also increased in COVID-19 Mild and Conv conditions compared to Cont. These results suggest a similarity in B-cell protective responses between vaccine and Mild/Conv conditions.

### Characterization of innate immune cells

To investigate vaccine (**Fig 6A-D**, **S6A**) and infection-driven (**Fig 6F**, **S6B-E**) changes in innate immune cells, the distribution and expression of classical subtype markers were used to classify innate cell types. We identified 6 innate cell types including NK cells, γδ T cells, MAIT cells, DC cells, monocytes and megakaryocytes (**Fig 6A-B, 6F, S6A-E**). The dendritic cells were further classified into 2 subtypes including pDCs, (plasmacytoid dendritic cells) and mDC (monocyte-derived dendritic cells), while monocytes were classified into 3 subtypes including CD16^+^ monocytes, CD14^+^ monocytes and CD14^+^CD16^+^ monocytes (**Fig 6A-B, 6F, S6A-E**).

**Fig 6.**
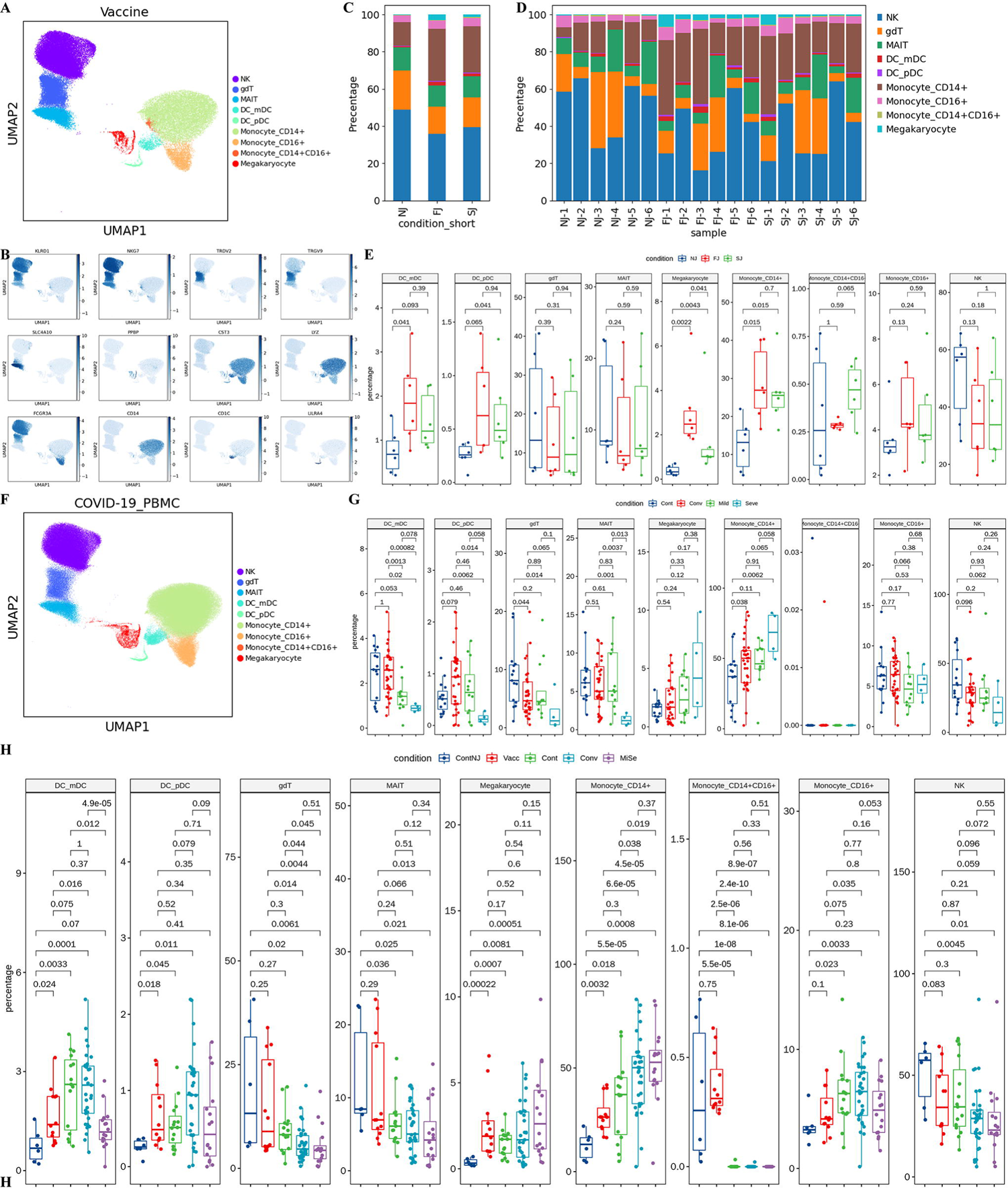
Characterization of innate cell composition differences in individuals across vaccination and infection conditions. **A.** UMAP projection of all innate cells from NJ, FJ and SJ conditions. Each dot corresponds to a single cell, colored by its cell subtype. **B.** Expression levels of canonical innate cell RNA markers were used to identify and label major cell clusters on the UMAP plot. Cells are colored according to log transformed and normalized expression levels of eight genes. Cells are from NJ, FJ and SJ conditions. **C.** Average proportion of each innate cell subtype derived from NJ, FJ and SJ groups. **D.** Proportion of each innate cell subtype derived from NJ, FJ and SJ individual samples. **E.** The box plot shows the composition of innate cells in NJ, FJ and SJ conditions at a single sample level. **F.** UMAP projection of all innate cells from Cont, Conv, Mild, and Seve conditions. Each dot corresponds to a single cell, colored by its cell subtype. **G.** Proportion of each innate cell subtype derived from Cont, Conv, Mild and Seve individual samples. **H.** Proportion of each innate cell subtype derived from ContNJ(NJ), Vacc(FJ+SJ), Conv, MiSe (Mild and Seve) individual samples. All pairwise differences with P < 0.05 using two-sided unpaired Mann–Whitney U-test are marked to show significance levels.

To obtain further insights into the features of innate cells, we examined the distribution of each subset across three conditions (NJ, FJ and SJ). The proportions of γδ T cells, NK cells, MAIT cells, CD16+ Mono, CD14+CD16+ Mono were similar across the three conditions (NJ, FJ and SJ) (**Fig 6E, 6H**, **S6F**), while the relative percentages of γδ T cells, NK cells and MAIT cells decreased after SARS-CoV-2 infection, especially in severe patients (**Fig 6G, 6H**, **S6F**). The innate cell subsets mDC, pDC, megakaryocytes and CD14^+^ monocytes increased after vaccination (**Fig 6E, 6H**, **S6F**). For COVID-19 patients, the percentages of mDC and pDC significantly decreased in severe patients but were restored in convalescent samples (**Fig 6G, 6H**, **S6F**). In contrast, the proportions of megakaryocytes and CD14^+^monocytes were significantly increased in severe patients and were also increased in convalescent samples (**Fig 6G, 6H**). These findings of changes to innate cell subsets in COVID-19 patients are consistent with previous reports and flow-cytometry-based results (Jouan et al., 2020; Wilk et al., 2020). These data also indicate that both vaccines and SARS-CoV-2 infection alter components of the innate cells in PBMCs and reveals distinct differences in innate cells between vaccine-induced immunity and natural SARS-CoV-2 infection.

### Transcriptomic changes in innate immune cells after vaccination and SARS-CoV-2 infection

Next, we investigated the transcriptomic changes in innate immune cells after vaccination. GO (Gene Ontology) analyses were conducted to obtain functional insights into innate cell subtypes between FJ/SJ conditions with NJ condition (**Fig 7A**). Genes associated with the adaptive immune response (such as “T cell activation”, “immune response-activating signal transduction” and “antigen processing and presentation”) were enriched after vaccination (**Fig 7A**), while for COVID-19 infection, other pathways (such as “response to virus” and “defense response to virus”) were enriched (**Fig S7A**). Genes associated with “Response to IFN signaling” were enriched in both vaccination and SARS-CoV-2 infection (**Fig 7A-B**, **S7B-7C**). INF response is an essential pathway for innate cells to respond to viral infections, and our data indicates that CoronaVac successfully induces the INF response in innate cells (**Fig 7B**, **S7B**). The “INF response” of innate immune cells was stronger in SARS-CoV-2 infection (especially for severe COVID-19 patients) compared to vaccination (**Fig 7B**). We observed that four innate immune cell types (monocytes, γδ T, MAIT and NK cells) exhibited significantly upregulated IFN after vaccination (**Figure S7B**), while all 6 innate cell types had higher IFN in COVID-19 patients (**Fig S7C**).

**Fig 7.**
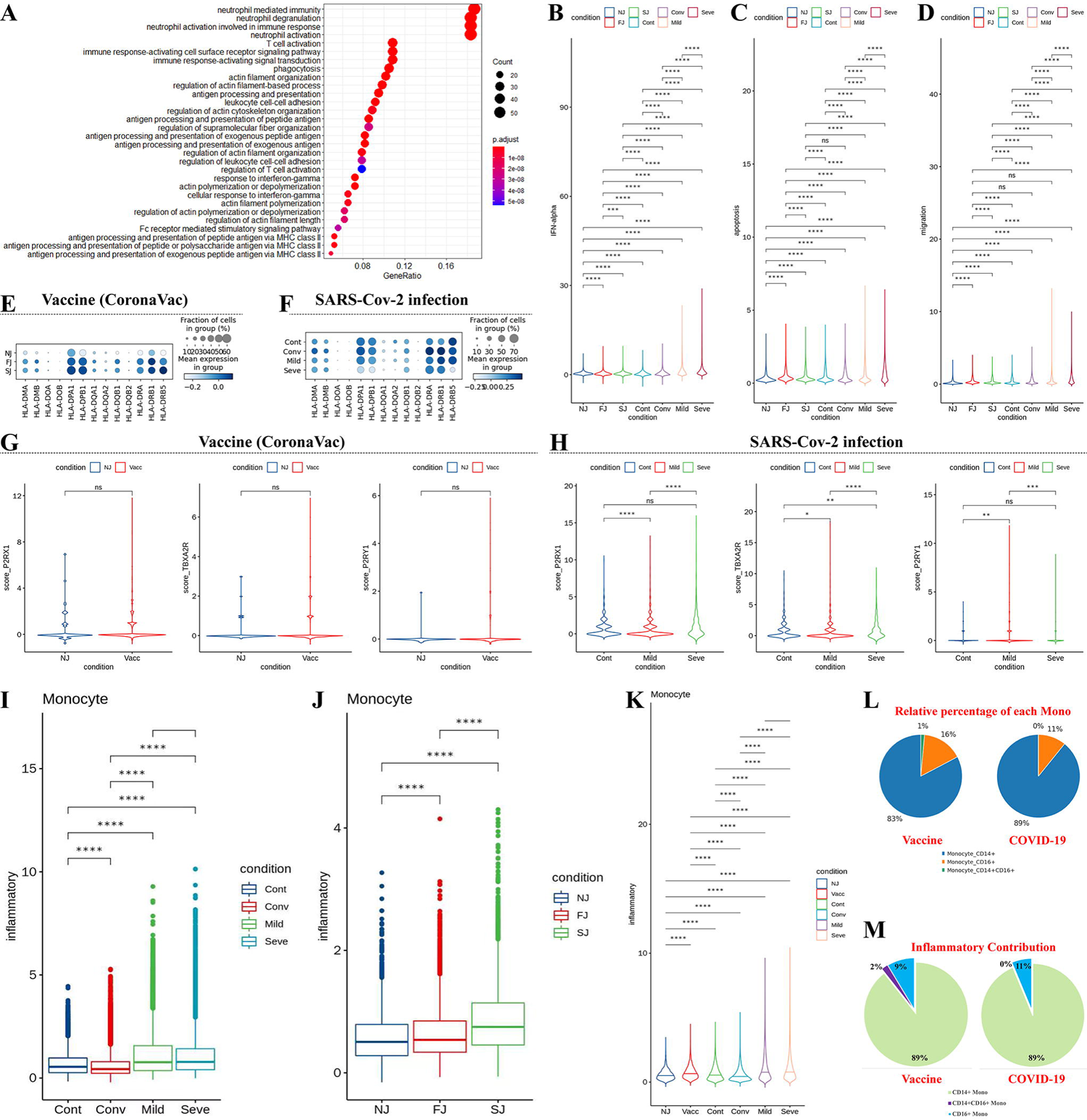
Characterization of gene expression differences in innate immune cells from vaccine and COVID-19 infected cohort samples. **A**, GO enrichment analysis of DEGs identified by comparing the before and after vaccination conditions. DEGs refer to genes with Benjamini–Hochberg adjusted P value (two-sided unpaired Mann–Whitney U-test) ≤0.01 and average log2 fold change ≥1 in both FJ/NJ and SJ/NJ comparisons. **B**, **C** and **D**, Expression activity of IFN-alpha, apoptosis and migration pathways in innate immune cells of NJ, FJ, SJ, Cont, Conv, Mild and Seve conditions shown as violin plots and colored by sample conditions. **E**, Heatmap dot plot of HLA-II gene expression in innate immune cells of NJ, FJ, and SJ conditions **F**, Heatmap dot plot of HLA-II gene expression in innate immune cells of Cont, Conv, Mild and Seve conditions **G**, Violin plot showing normalized expression levels of *P2RX1*, *TBXA2R* and *P2RY1* in megakaryocytes (Mega) from NJ and Vaccine (FJ+SJ) conditions **H**, Violin plot of normalized expression of *P2RX1*, *TBXA2R* and *P2RY1* in megakaryocytes (Mega) from Cont, Mild and Seve conditions **I**, Expression activity of inflammatory pathways in monocytes from NJ, FJ and SJ conditions shown as box plots. Boxes are colored by sample conditions. **J**, Expression activity of inflammatory pathways in monocytes from Cont, Conv, Mild and Seve conditions shown as box plots. Boxes are colored by sample conditions. **K**, Expression activity of inflammatory pathways in monocytes from NJ, Vacc, Cont, Conv, Mild and Seve conditions shown as violin plots. Violins are colored by sample conditions. **L**, Pie graph of relative percentage for CD14+monocytes, CD16+monocytes, CD14+CD16+monocytes in the vaccine and COVID-19 cohort. **M**, Pie graph of inflammatory scores from CD14+monocytes, CD16+monocytes, CD14+CD16+monocytes in the vaccine and COVID-19 cohort.

Our data showed that levels of cellular apoptosis and migration were significantly upregulated in innate cells at the bulk level after vaccination and SARS-CoV-2 infection (**Fig 7C, S7D-E**). The expression of HLA-II genes at FJ and SJ conditions was higher compared to NJ, suggesting the enhancement of crosstalk across cells after vaccination (**Fig 7E**). However, some HLA-II genes (e.g., *HLA-DRB5*, *HLA-DPB1* and *HLA-DPA1*) were downregulated after SARS-CoV-2 infection, especially in severe COVID-19 patients (**Fig 7F**), implying possible impairment of crosstalk across cells. Genes which encoded HLA class I molecules (such as HLA-A, HLA-B and HLA-E) were upregulated in innate cells from immunized samples relative to NJ condition (**Fig S8**). Similar results were also seen in COVID-19 patients (**Fig S9**). The underlying mechanism and effect of changes in HLA-I molecules requires further investigation. In addition, we also investigated the expression of several critical genes related with platelet aggregation (P2RX1, P2RY1 and TBXA2R) in megakaryocytes (Sangkuhl et al., 2011). P2RX1, P2RY1 and TBXA2R were not significantly upregulated after immunization which suggests a low risk of thrombosis following CoronaVac immunization (**Fig 7G**). P2RX1, P2RY1 and TBXA2R were significantly upregulated in mild COVID-19 patients (**Fig 7G**), which may imply a higher risk of thrombosis for mild COVID-19 patients than severe COVID-19 patients.

We further analyzed monocytes, as previous reports suggested that this cell subset appeared to be the source of inflammation in COVID-19 patients (Ren et al., 2021). We evaluated the expression of genes reported to encode inflammatory cytokines (**Table S3**) (Ren et al., 2021; Wilk et al., 2020). We found elevated expression of inflammatory genes in COVID-19 patients compared to healthy controls at the bulk level, indicating that peripheral monocytes are potential contributors to the inflammatory cytokine storm observed in COVID-19 patients. However, severe COVID-19 patients did not show higher expression of inflammatory cytokines compared to mild patients (**Fig I**). We also identified increased expression of inflammatory response genes in vaccinated individuals (FJ and NJ conditions) compared to NJ condition at the bulk level, especially after the second dose (**Fig 7J**). Of note, expression of inflammatory response genes in vaccinated individuals was much lower than COVID-19 patients (**Fig K**). This implies that post-vaccination may not cause an increase in inflammatory cytokines in peripheral blood or causes a lower increase in inflammatory cytokines. To validate this result, we investigated the levels of 11 cytokines (including pro-inflammatory cytokines: TNF-α, IL-1B and IL-6) in the sera of the 18 samples using a bead-based flow-cytometry assay on the BD LSRFortessa X-20 platform. No obvious post-vaccination elevation in most cytokines were observed (**Fig S10**), which further suggests that post-vaccination does not lead to significant increases in inflammatory cytokines in peripheral blood. Interestingly, CD14+monocytes contributed to the highest proportion of cell composition (**Fig 7L**) and inflammatory scores (**Fig 7M**) after vaccination or SARS-CoV-2 infection, suggesting that CD14+ monocytes may be the major source of inflammation (Ren et al., 2021; Zhou et al., 2020).

### Features of T cell subsets in individuals after vaccination and SARS-CoV-2 infection

To investigate changes in individual T cell subclusters, the T cells from PBMCs of vaccinated individuals across three conditions (NJ, FJ and SJ) (**Fig 8A-B**, **S11A**) and in COVID-19 patients (**Fig S11B-D**) were subclustered into 12 subtypes according to the distribution and expression of classical T cell markers. These include 7 CD4^+^T cell subtypes (CD3D^+^ CD3E^+^ CD40LG) and 5 CD8^+^T cell subtypes (CD3D^+^ CD3E^+^ CD8A^+^CD8B^+^).

**Fig 8.**
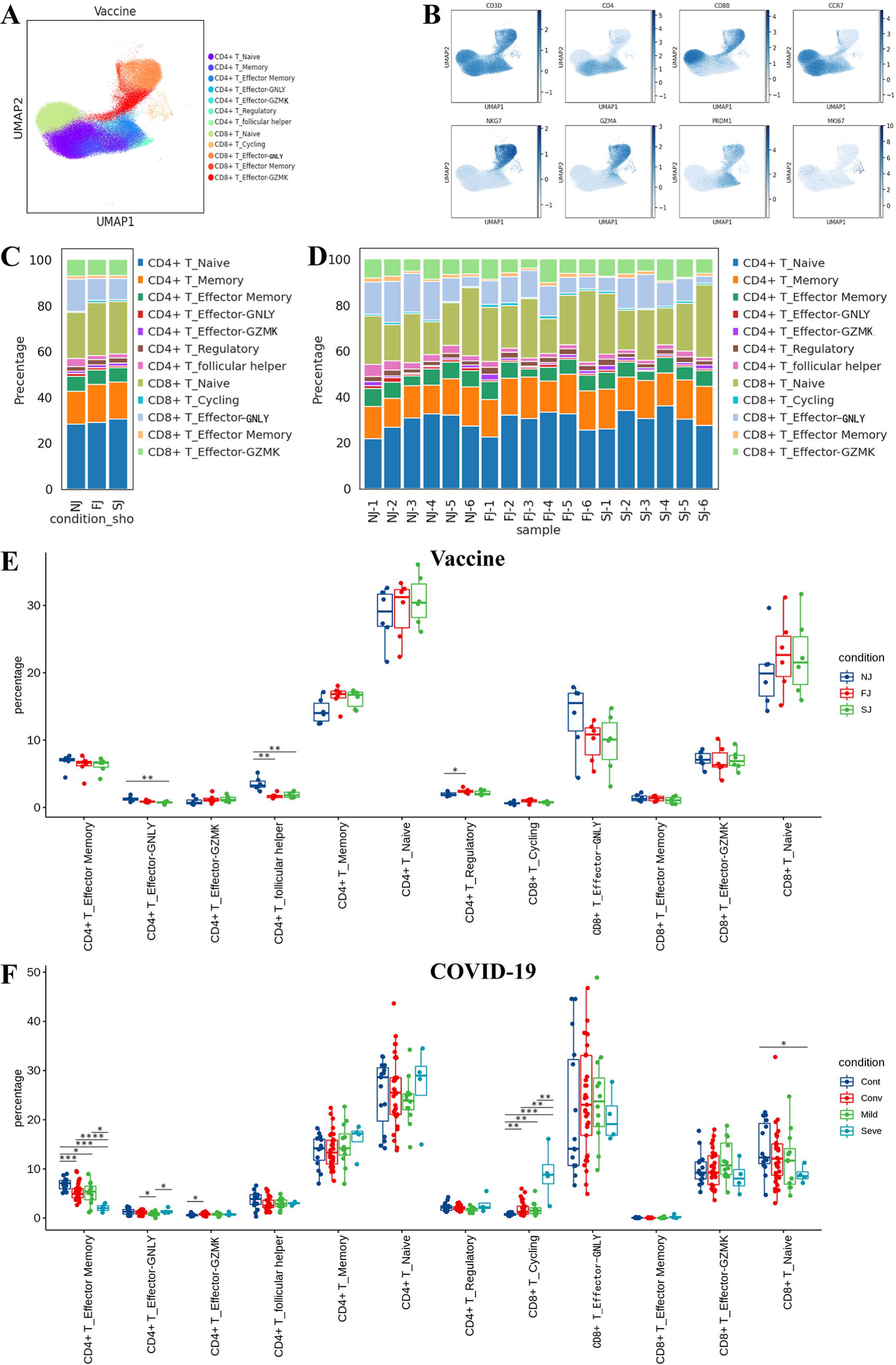
Characterization of T cell composition differences in individuals across vaccination and infection conditions. **A**, UMAP projection of all T cells from NJ, FJ and SJ conditions. Each dot corresponds to a single cell, colored by its cell subtype. **B**, Expression levels of canonical T cell RNA markers were used to identify and label major cell clusters on the UMAP plot. Cells are colored according to log transformed and normalized expression levels of eight genes. Cells are from NJ, FJ and SJ conditions **C**, Average proportion of each T cell subtype derived from NJ, FJ and SJ groups. **D**, Proportion of each T cell subtype derived from NJ, FJ and SJ individual samples. **E**, The box plot shows the composition of T cells from NJ, FJ and SJ conditions at a single sample level. **F**, The box plot shows the composition of T cells from Cont, Conv, Mild and Seve conditions at a single sample level. All pairwise differences with P < 0.05 using two-sided unpaired Mann–Whitney U-test are marked to show significance levels.

For CD4^+^T cells, we defined one naïve CD4^+^ T cell subset (CCR7+SELL+), one memory CD4^+^ T cell subset (S100A4^+^GPR183^+^), one effector memory CD4^+^ T cell subset (S100A4^+^GPR183^+^GZMA^+^), one regulatory CD4^+^ T cell subset (T_reg_; FOXP3^+^IL2RA^+^), one follicular T helper (Tfh) cell subset (CXCR5^+^, ICOS+, SLAMF1+) and two effector CD4^+^ T cell subsets (CD4^+^ effector-GZMK and CD4^+^ effector-GNLY). Notably, the CD4+ effector-GNLY cell subtype was characterised with high expression of genes related with cytotoxicity, such as *GNLY*, *GZMB*, *NKG7* and *KLRD1*, whereas the CD4^+^ effector-GZMK cell subtype displayed high expression of *GZMK* and low expression of other cytotoxic genes (**Fig 8A-B**, **S11A, Table S5**). The CD4+ effector-GNLY subtype also had highly expressed TBX-21 (T-bet), suggesting that this subcluster were Th-1 (Type 1 helper)-like cells (**Table S5**). For CD8^+^ T cells, we defined one naïve CD8^+^ T cell subset (CCR7^+^SELL^+^), one cycling CD8+ T cell subset (MKI67+), one effector memory CD8^+^ T cell subset (S100A4^+^GPR183^+^GZMA^+^), two effector CD8^+^ T cell subsets (CD8^+^ effector-GNLY and CD8^+^ effector-GZMK) (**Fig 8A-B**, **S11A, Table S5**).

To gain further insights into the characteristics of the T cell subclusters, we examined the distribution of each subtype across three vaccine timepoints (**Fig 8C-E**) and compared these profiles with data collected from COVID-19 patients (**Fig 8F**, **S11E-G**). Three T cell subsets were significantly altered after vaccination (**Fig 8E**) in comparison to NJ with CD4+ effector-GNLY and Tfh cell subsets decreasing while Treg cells increased. For SARS-CoV-2 infection, the relative percentage of naïve CD8+ T cells significantly decreased in severe COVID-19 patients whereas no significant changes were observed in naïve CD4+ T cell subsets (**Fig 8F**). The proportion of CD4+ effector memory decreased in COVID-19 patients compared to controls (**Fig 8F**) and in the convalescence stage, the CD4+ effector memory cells remained low and were not restored to the levels observed in the controls. In contrast, two cytotoxic subsets, including CD4+ effector-GNLY and CD4+ effector-GZMK, were present in higher percentages for convalescence patients (**Fig 8F**). Of particular note, the cycling CD8+ subset was almost absent in controls but were highly enriched in COVID-19 patients, especially for severe patients (**Fig 8F**). Besides naïve CD8+, CD4+ effector-GNLY, CD4+ effector-GZMK, CD4+ effector memory and cycling CD8+ subsets, others T cell subsets were not significantly altered (**Fig 8F**, **S11F-G**).

### Transcriptomic changes in T cells after vaccine and SARS-CoV-2 infection

We investigated transcriptomic changes in the T cell subsets of vaccinated participants and identified differences between vaccine and natural infection induced responses. GO analyses found that genes associated with “T cell activation”, “antigen processing and presentation” and “response to interferon” were enriched in T cell subsets after vaccination, implying an ongoing adaptive immune response to vaccination (**Fig 9A**). For SARS-CoV-2 infection, “Interferon signaling pathway”, “response to virus” and “defense response to virus” were specifically enriched in T cell subsets, suggesting an ongoing response against the virus (**Fig S12A**). IFN response is essential to the immune response triggered by vaccines or viral infections and consistently, we found that T cell subsets exhibited significant upregulation of IFN after vaccination and SARS-CoV-2 infection (**Fig 9B**, **S12B**). Four activated state T cell subsets, including CD4+ effector memory, CD4+ effector-GZMK, CD8+ effector-GNLY and CD8+ effector memory, showed significantly upregulated IFN after vaccination, whereas in SARS-CoV-2 infection, all activated state T cell subsets had IFN significantly upregulated (**Fig 9C**, **S12B**). COVID-19 patients had stronger expression of IFN than vaccinated individuals (**Fig 9D**).

**Fig 9.**
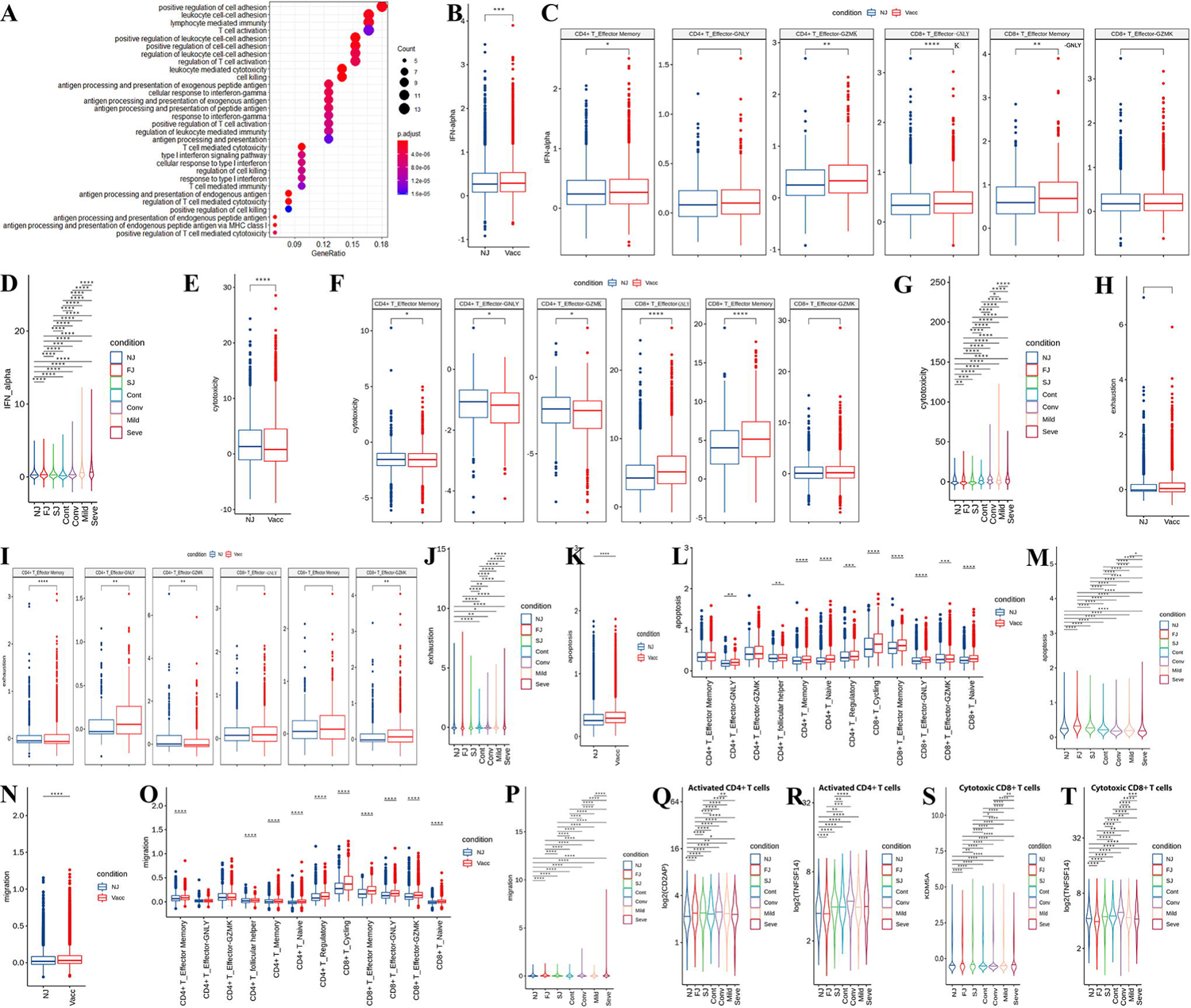
Characterization of gene expression differences in activated T cells from vaccine and COVID-19 infected cohort samples. **A**, GO enrichment analysis of DEGs identified by comparing the before and after vaccination conditions. DEGs refer to genes with Benjamini–Hochberg adjusted P value (two-sided unpaired Mann–Whitney U-test) ≤0.01 and average log2 fold change ≥1 in both FJ/NJ and SJ/NJ comparisons. **B** and **C**, Expression activity of IFN-alpha pathways in activated T cells (**B**) and subtypes (**C**) of NJ and Vacc (FJ and SJ) conditions shown as box plots and are colored by sample conditions. **D**, Expression activity of IFN-alpha in activated T cells of NJ, FJ, SJ, Cont, Conv, Mild and Seve conditions shown as violin plots and colored by sample conditions. **E** and **F**, Expression activity of cytotoxicity pathways in activated T cells (**E**) and subtypes (**F**) of NJ and Vacc (FJ and SJ) conditions shown as box plots and are colored by sample conditions. **G**, Expression activity of cytotoxicity pathways in activated T cells of NJ, FJ, SJ, Cont, Conv, Mild and Seve conditions shown as violin plots and colored by sample conditions. **H** and **I**, Expression activity of exhaustion genes in activated T cells (**H**) and subtypes (**I**) of NJ and Vacc (FJ and SJ) conditions shown as box plots and are colored by sample conditions. **J**, Expression activity of exhaustion genes in activated T cells of NJ, FJ, SJ, Cont, Conv, Mild and Seve conditions shown as violin plots and colored by sample conditions. **K** and **L**, Expression activity of apoptosis pathways in T cells (**K**) and subtypes (**L**) of NJ and Vacc (FJ and SJ) conditions shown as box plots and are colored by sample conditions. **M**, Expression activity of apoptosis pathways in activated T cells of NJ, FJ, SJ, Cont, Conv, Mild and Seve conditions shown as violin plots and colored by sample conditions. **N** and **O**, Expression activity of migration pathways in activated T cells (**N**) and subtypes (**O**) of NJ and Vacc (FJ and SJ) conditions shown as box plots and are colored by sample conditions. **P**, Expression activity of migration pathways in activated T cells of NJ, FJ, SJ, Cont, Conv, Mild and Seve conditions shown as violin plots and colored by sample conditions. **Q** and **R**, Gene expression level of CD2AP (**Q**) and TNFSF14 (**R**) in activated CD4+T cells in NJ, FJ, SJ, Cont, Conv, Mild and Seve conditions. Violin plots showed normalized average expression of CD2AP (**Q**) and TNFSF14 (**R**). **S** and **T**, Gene expression level of KDM5A (**S**) and TNFSF14 (**T**) in cytotoxic CD8+T cells in NJ, FJ, SJ, Cont, Conv, Mild and Seve conditions. Violin plots showed normalized average expression of KDM5A (**S**) and TNFSF14 (**T**). All pairwise differences with P < 0.05 using two-sided unpaired Mann–Whitney U-test are marked to show significance levels.

We then evaluated the cytotoxicity scores of different effector T cell subsets after vaccination. Our data showed that effector T cell subsets had lower cytotoxicity scores after vaccination than NJ condition at the bulk level (**Fig 9E**), and only two effector T cell subsets (CD8+ effector-GNLY and CD8+ effector memory) showed higher cytotoxicity scores after vaccination (**Fig 9F**). In contrast, the effector T cell subsets exhibited higher cytotoxicity scores in COVID-19 patients than controls at the bulk level, and all effector T cell subsets showed higher cytotoxicity scores in COVID-19 patients (**Fig S12C**). COVID-19 patients also exhibited higher cytotoxicity scores than vaccinated individuals (**Fig 9G**). Interestingly, we did not observe significant elevation in exhaustion scores for effector T cell subsets after vaccination at the bulk level (**Fig 9H**) however only CD4+ effector-GNLY and CD8+ effector-GZMK showed an increase in exhaustion scores after vaccination (**Fig 9I**). For SARS-CoV-2 infection, effector T cell subsets did show higher exhaustion scores at the bulk level (**Fig S12D**). All effector T cell clusters in mild and severe COVID-19 patients had higher exhaustion scores than controls, with severe patients having the highest exhausted score (**Fig 9J, S12D**). COVID-19 infection did not displayed a stronger exhaustion score in comparison to vaccinated individuals (**Fig 9J**).

We also investigated the apoptosis and migration scores of different T cell subsets. Our data indicated that both vaccination and SARS-CoV-2 infection showed a high level of apoptosis and migration in T cell subsets at the bulk level (**Fig 9K-P**, **S12E-F**), however stronger apoptosis and migration of T cell subsets were observed in COVID-19 patients, especially in the severe group (**Fig 9K-P**, **S12E-F**). These results suggest that significant activation of cell apoptosis and migration pathways in the PBMCs of severe disease may be associated with lymphopenia, consistent with previous studies (Chen et al., 2020; Tan et al., 2020).

In addition, we observed that the expression of CD2AP significantly upregulated on activated CD4+ T cells after vaccination and SARS-CoV-2 infection (**Fig 9Q**). CD2AP, as an adaptor protein in CD4+ T cell, can modulate the differentiation of Tfh cells and promote protective antibody responses in viral infection (Raju et al., 2018). The expression of CD258 (TNFSF14) also significantly upregulated on activated CD4+ T cells (**Fig 9R**) and cytotoxic CD8+ T cells (**Fig 9T**) after vaccination and SARS-CoV-2 infection. TNFSF14 serves as a key component for T cell recruitment to tissues from peripheral blood as well as promote T cell activation. The expression of KDM5A significantly elevated in cytotoxic CD8+ T cells after vaccination and SARS-CoV-2 infection (**Fig 9S**). KDM5A encodes a demethylase-H3K4me3, which is needed for T cell activation. These data suggested that increased activation of T cells in vaccinated individuals and COVID-19 patients may contribute to defense against the vaccine and SARS-CoV-2 virus.

### V(D)J gene usage and clonal expansion in T cells after vaccination and SARS-CoV-2 infection

TCR information was detected in all subsets and in ∼60% of T cells. Clonal expansion of CD8+ effector T cells was larger (clonal size >100) than CD8+ cycling T cells and CD4+ T cells (**Fig10A-B**). >70% of T cells in the vaccine cohort and 53%∼61% of T cells in COVID-19 cohorts had unique TCR clonotypes (**FigS13A-B**). A negative correlation between clone size and clonotype number was observed, consistent with previous report (Zhang et al., 2020) and suggests that large clonal expansion is a rare event (**Fig 10C**). Next, we compared the expression of T cell receptor β-chain constant domains 1 and 2 (TRBC1 and TRBC2). The percentage of TRBC1 increased after vaccination, while in COVID-19 patients, the percentage of TRBC1 was highest in severe disease and lowest in controls (**Fig 10D, H, S13C**). This suggests that immune activation by vaccination and infection are able to modulate T cell receptor β-chain ratios.

**Fig 10.**
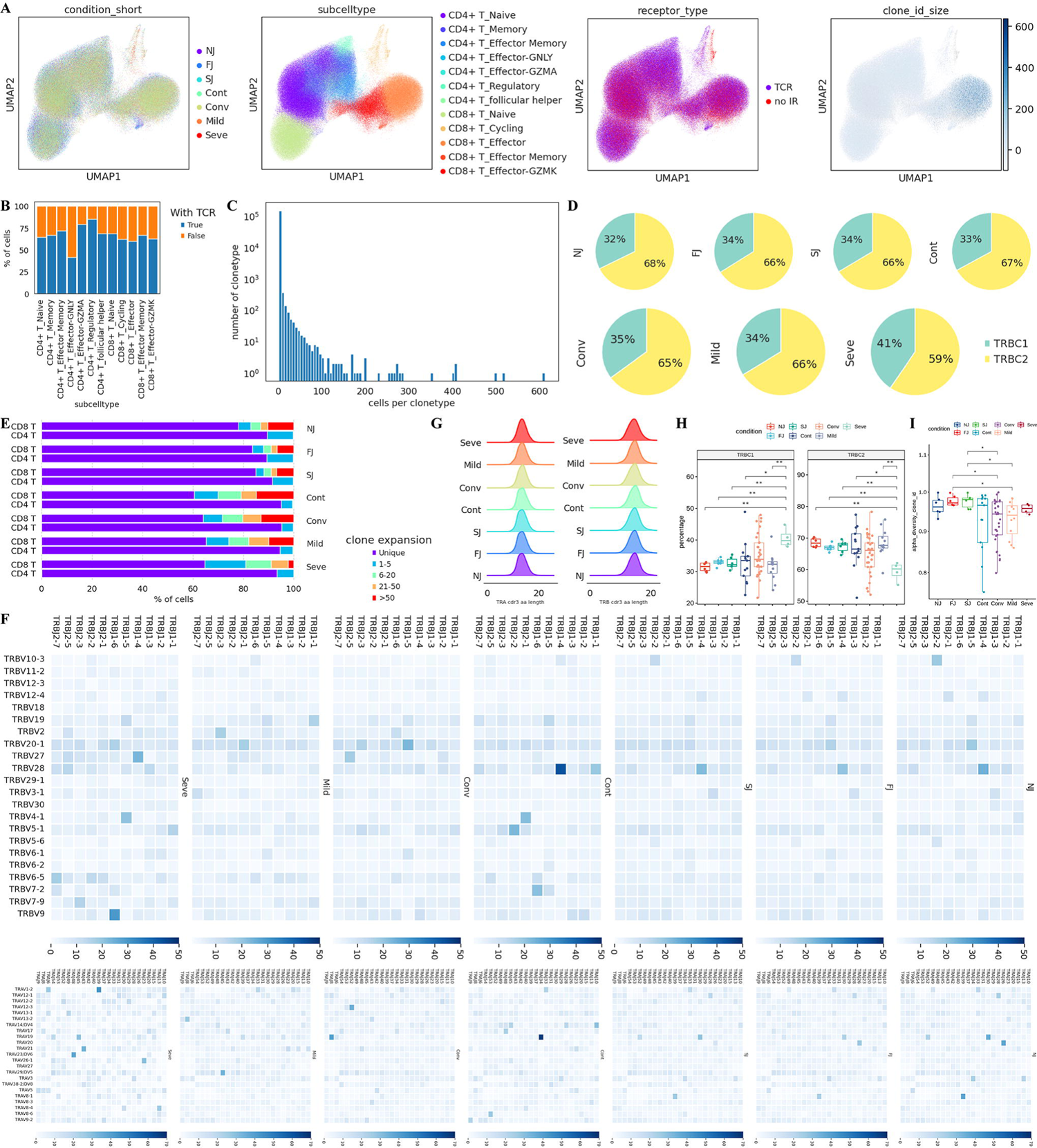
Changes in TCR clones and selective usage of V(D)J genes. **A**, UMAP projection of T cells derived from PBMCs. Cells are colored by conditions (Panel 1), T cell subtypes (Panel 2), if TCR detection was successful (Panel 3) and clonotype expansion size (panel 4). **B**, Stacked bar plot shows the TCR detection success rate for each T cell subtype. **C**, Histogram shows the negative correlation between the number of T cell clones and the number of cells per clonotype. Y axis is log10 scaled. **D**, Pie graph showing the distribution of TRBC1 and TRBC2 in T cells under each condition. **E**, Stacked bar plots showing the clone state of each T cell subtype in each condition. **F**, Heat maps showing differential TRBV-J and TRAV-J rearrangement. Prevalent TRBV-J combination pairs (top) and TRAV-J combination pairs (bottom) are compared across conditions. Usage percentage are sum normalized by column. **G**, Density curve plots showing the distribution shift of TRA and TRB chain CDR3 region length in TCR clone types from each condition. **H**, Box plot showing TRBC1 and TRBC2 percentages in NJ, FJ, SJ, Cont, Conv, Mild and Seve conditions **I**, Box plot showing the alpha diversity of TCR clonotypes in each PBMC sample. Data points are colored by condition.

We compared T cell clonal expansion after vaccination and observed a significant decrease in clonal expansion of CD8+ T cells, especially CD8+T effector-GNLY cells, from NJ to FJ to SJ. This suggests that two-doses of CoronaVac induced immunogenic proliferation leading to many new unique TCR clonotypes (**Fig 10E**, **S13D)**. For COVID-19 infection, decreased CD8+ T cell clonal expansion was also observed in infected patients compared to Cont and Conv (**Fig 10E**), in agreement with previous report (Tanriover et al., 2021). Clonal expansion in CD4+ T cells was also decreased from NJ to SJ but was higher in severe than controls, suggesting that CD4+ T cells may play different roles in vaccine and infection immune responses (**Fig 10E**).

The length distribution of the CDR3 region were similar for all conditions (**Fig10G**). COVID-19 infected patients tended to have lower TCR diversity while vaccinated patients had higher diversity (**Fig 10I**) which may suggest a protective role for TCR diversity. The usage of TRB V(D)J genes across vaccine and infection conditions were compared (**Fig 10F**). The combination of the most prevalent 11 TRBJ and 22 TRBV genes indicates that vaccination induced greater diversity in TRB V(D)J genes (**Fig10F** top panel). Mild COVID-19 infection also induced greater diversity however no new V(D)J pair patterns were observed. In contrast, severe COVID-19 infection induced new prominent V(D)J pairs, including TRBJ1-6/TRBV9, TRBJ1-4/TRBV27 and TRBJ1-5/TRBV4-1. The change in pattern for TRAJ and TRAV pairs showed similar trends to TRB V(D)J (**Fig10F** bottom panel). In addition, the CDR3 peptides sequences showed large individual differences however we did not observe any interesting patterns across conditions (**Fig S14**).

## Discussion

As an emerging virus, SARS-CoV-2 is highly pathogenic and is responsible for the COVID-19 pandemic. Currently, there are no effective drugs or optimal treatments for SARS-CoV-2 infection, thus considerable effort has been put into developing safe and effective vaccines against COVID-19. Inactivated SARS-CoV-2-based vaccines are one of the most-widely used COVID-19 vaccines due to its low cost, ease of scale-up and production (Iversen and Bavari, 2021a). CoronaVac is an inactivated COVID-19 vaccine candidate which has had its safety and potency validated in both animal models and clinical trials (Tanriover et al., 2021; Zhang et al., 2021). However, current knowledge of the host immune response to the inactivated COVID-19 vaccine is still limited, making it difficult to inform and improve the design of new generations of COVID-19 vaccines. In addition, little is known about how this immune response compares to natural SARS-CoV-2 infection.

In this report, we performed single-cell RNA sequencing in PBMCs of six individuals immunized with CoronaVac and compared these to the single-cell profiles in COVID-19 patients. Overall, both inactivated SARS-CoV-2 vaccine and natural SARS-CoV-2 infection affect the composition of peripheral immune cells, with greater changes observed in COVID-19 patients, especially for severe disease. After CoronaVac injection, the inactivated SARS-CoV-2 virus initially encounters antigen presenting cells (APCs) in the innate immune system which then triggers the adaptive immune response. Our data confirmed that the proportion of DCs, one of the main APCs, are significantly elevated after vaccination (**Fig 2**), thus suggesting that CoronaVac is able to successfully initiate the adaptive immune response. However in severe COVID-19 patients, the relative percentage of DCs significantly decreased, implying a possible subversion of the adaptive immune response in severe disease. Monocytes are the major source of inflammatory cytokines in SARS-CoV-2 infection (Ren et al., 2021; Zhou et al., 2020), and were elevated after immunization with CoronaVac and SARS-CoV-2 infection (**Fig S2**). However, the level of monocytes was significantly greater in COVID-19 patients (especially for severe disease) compared to immunization. This implies that CoronaVac immunization causes a weaker inflammatory response than natural SARS-CoV-2 infection. The relative abundance of γδ T, MAIT, NK, CD4+ effector memory, and naïve CD8+ T cells decreased with disease severity while the proportion of cycling CD8+ T cells increased with disease severity, suggesting that these subsets may be associated with disease severity. We did not observe significant changes to these subsets after immunization with CoronaVac (**Fig 2, 6, 9, S2, S6** and **S11**).

IFNs are produced during viral infection and has an antiviral role. However when produced excessively, IFNs can cause immunopathological damages. Interestingly, ‘IFN response’ was enriched by GO analysis in different cell subclusters after vaccination in our study (**Fig 4, 7 and 9**). Compared to pre-vaccination samples, the response to IFN-α (type I IFN) pathway was also significantly elevated in most PBMC cell types in post-vaccination samples. Together, this suggests CoronaVac induces IFNs as part of the antiviral response. Similarly, ‘IFN response’ was also enriched by GO analysis after SARS-CoV-2 infection (**Fig S4, S7** and **S12**). The expression level of IFN-α was also significantly upregulated in COVID-19 patients (**Fig S4, S7** and **S12**), suggesting an antiviral role triggered by IFNs. Notably, COVID-19 patients showed higher response to IFN-α than immunized individuals with the strongest response to IFN-α found in severe disease (**Fig 7, 9**).This indicates that the stronger response to IFNs in COVID-19 patients may be involved in immune pathology (e.g., lung injury) (Liu et al., 2020).

In this report, we observed broad immune activation after immunization with CoronaVac. First, the levels of neutralizing antibodies (anti-S-RDB-specific antibody) significantly increased after the second injection (**Fig 3**, **S3**, **Table S2**). Second, B cell activation in PBMCs post-vaccination is supported by our data and evidenced by: (i) enrichment of genes involving in ‘B cell activation’, ‘adaptive immune response’ and ‘antigen processing and presentation’; (ii) significant upregulation of two important B cell activation pathways (GO:0002312 and GO:0042113); (iii) the activation of naïve B and memory B cell subsets; (iv) significant expression of key genes (e.g., *PRDM1*, *XBP1*, *IFR4*, *PAX5*, *IL4R* and *IL21R*). Third, the activation of innate immune cells was observed in our study as genes associated with ‘antigen processing and presentation’, ‘IFNs response’, ‘T cell activation’ and ‘immune response-activating signal transduction’ were enriched by GO analysis (**Fig 7**). Fourth, the activation of T cell subsets was also supported by our data as genes involved in ‘T cell activation’, ‘T cell mediated immunity’, ‘antigen processing and presentation’, and ‘IFNs response’ were also enriched by GO analysis (**Fig 9**). Similarly, broad immune activation was also observed after SARS-CoV-2 infection (**Fig S4, S7, S12**). However, we observed significant downregulation of some HLA class II genes in B, T and innate cells in COVID-19 patients, especially for severe disease, implying a dysregulation in crosstalk between adaptive immune cell classes. Furthermore, some key chemokine receptor genes (e.g., CXCR5) were also significantly downregulated in the PBMCs of severe COVID-19 patients (**Fig S4**), which may impair germinal center reaction, resulting in a dysregulated humoral immune response (Mathew et al., 2020).

We analyzed the apoptosis, migration, cytotoxicity and exhaustion scores in different immune cell subsets from immunized individuals and compared their expression levels with COVID-19 patients. Overall, innate immune and T cell subsets showed higher apoptosis and migration scores after immunization with CoranaVac or SARS-CoV-2 infection (**Fig 7, 9, Fig S7** and **S12**). However, compared to post-vaccination, SARS-CoV-2 infection exhibited higher apoptosis and migration scores, with severe disease having the highest score. This suggests that severe patients likely had increased lymphocyte apoptosis and migration which may be associated with lymphopenia, a clinical predictor for severe COVID-19 disease (Tan et al., 2020). At the bulk level, post-vaccination effector T cell subsets did not display higher exhaustion scores compared to pre-vaccination samples. However for COVID-19 patients, all samples had higher exhaustion scores compared to controls, and those with severe disease displayed the highest exhaustion scores. It is possible that the high exhaustion status of effector T cell subsets may be associated with functional impairment (Zheng et al., 2020). Interestingly, the effector T cell subset from post-vaccination showed lower cytotoxicity scores at the bulk level than those of pre-vaccination. In contrast, the cytotoxicity scores of the effector T cell subset from COVID-19 patients were higher than the controls with severe disease having the highest cytotoxic score. Similarly, we observed that the cytotoxicity scores in each subset of effector T cells were also significant elevated after SARS-CoV-2 infection.

Although increased expression of pro-inflammatory cytokine genes was observed in monocytes post-vaccination, this upregulation may not be adequate to cause a significant increase in systemic levels of pro-inflammatory cytokines. This is supported by our immunoassay results which showed no obvious increase in several key pro-inflammatory cytokines (e.g., IL-6 and TNF) post-vaccination (**Fig S10**). In addition, the expression level of pro-inflammatory cytokine genes after vaccination was significantly lower than SARS-CoV-2 infection. These evidences suggest that CoronaVac may not contribute to acute inflammation or the cytokine storm commonly observed in severe COVID-19 patients (Zhou et al., 2020). Among the monocyte subsets, CD14+ monocytes were identified as the major contributor to inflammation for both immunization and SARS-CoV-2 infection. This is evidenced by: (i) a large increase in CD14+ monocytes observed after vaccination and SARS-CoV-2 infection (**Fig 3 and Fig S3**) (Zhou et al., 2020) and (ii) CD14+ monocytes was the largest contributor to the inflammatory scores (**Fig 7**). In addition, due to rising concerns about the bloodclotting side effect observed in several vaccine types, we also examined several key genes (e.g., *2RX1*, *P2RY1* and *TBXA2R*) involved in platelet aggregation in megakaryocytes (Sangkuhl et al., 2011). Unlike SARS-CoV-2 infection, our data demonstrated that the expression level of platelet aggregation-associated genes were not significantly upregulated after immunization with CoronaVac. This may explain the fewer number of thrombus-related adverse events reported CoronaVac.

### Methods Volunteer cohort

Three male and three female healthy adults (n = 6) were admitted at the Sanya People’s Hospital and enrolled in the study. Peripheral blood samples were collected at 3 key timepoints (Fig. 1A): pre-vaccine baseline (timepoint 1, day 0), 3 weeks following the first dose (timepoint 2, day 21), which was also the same day they received the second dose, and finally, 2 weeks following the second dose (timepoint 3, day 35). All six volunteers had matching samples at 3 time points. This study design allowed us to investigate the kinetics of the immune responses following both primary and secondary immunizations.

#### Sample collection

**Supplementary Table 1** summarizes the characteristics of individuals assessed in each assay. Full cohort information is described in figure 1. The fresh blood samples for each timepoint immediately underwent peripheral blood mononuclear cells (PBMCs) isolation using standard density gradient centrifugation. PBMCs are typically employed to assess immune-regulatory effects at the single-cell level. PBMCs were isolated using HISTOPAQUE-1077 (Sigma-Aldrich, 10771) solution according to the manufacturer’s instructions. Briefly, 3 mL of fresh peripheral blood was collected in EDTA anticoagulant tubes and subsequently layered onto HISTOPAQUE-1077. After centrifugation, PBMCs remained at the plasma-HISTOPAQUE-1077 interface and were carefully transferred to a new tube. Erythrocytes were removed using Red Blood Cell Lysis Solution (10×) (Miltenyi, 130-094-183) and washed twice with 1× PBS (Gibco, 10010023). The cell pellets were re-suspended in sorting buffer (PBS supplemented with 2% fetal bovine serum (FBS, Gibco, 10099141)). Cell viability of PBMCs were assessed using the Countstar cell viability detection kit and showed greater than 90% viability. PBMCs were then used in immunological analysis and cell encapsulation. The 10x Genomics single-cell transcriptome platform was used to generate the 5’ gene expression profiles, T cell receptor (TCR) and B cell receptor (BCR) data. This approach employs a commercial emulsion-based microfluidic platform (Chromium 10x) that enables the generation of amplified cDNA used for both the preparation of single cell RNA-seq libraries and TCR/BCR target enrichment and sequencing.

#### Single cell RNA library preparation and sequencing

Cell suspensions were barcoded through the 10x Chromium Single Cell platform using Chromium Single Cell 5’ Library, Gel Bead and Multiplex Kit, and Chip Kit (10x Genomics). Single-cell RNA libraries were prepared using the Chromium Single Cell 5’ Kit v2 (10x Genomics; PN-1000263), Chromium Single Cell V(D)J Reagent kits (10x Genomics, PN-1000252(TCR), PN-1000253(BCR)) according to the manufacturer’s instructions. Each sequencing library was generated with a unique sample index. The libraries were sequenced on the Illumina Novaseq6000 sequencer with a paired-end 150-bp (PE150) reading strategy. With the provided sample sheet, the CellRanger (v.5.0.0) *mkfastq* command was used to demultiplex the flow cells’ raw base call files into fastq files.

#### SARS-CoV-2-specific IgM/IgG ELISA and plasma cytokine detection

The S-specific IgG/IgM and plasma cytokine detection were detected according to our previous study (Wang et al., 2021).

### Quantification and Statistical analysis Single-cell RNA-seq and data processing

The human reference (v.GRCh38-3.0.0) was downloaded from the 10x Genomics official website in Mar-2021. Raw and filtered gene expression matrices were generated for each sample using the kallisto/bustools (kb v0.24.4) pipeline coupled with human GRCh38. The *kb count* command was called to generate single cell feature counts for each sample by specifying the library name in the argument. Then the filtered feature, barcode and matrix files were analyzed in python (v3.8.10) using the anndata (ad) (v0.7.6) and scanpy (sc) (v1.7.2) packages. Data files of all 18 samples and the largest dataset of Chinese PBMC COVID-19 infection (64 fresh PBMC samples from Cell 2021 Zhangzemin) were merged together by the ad.concat function. Cells and genes were filtered by the sc.pp.filter_cells and sc.pp.filter_genes function for further analyses. First, genes expressed at a proportion >0.1% of the cells were selected. Second, to minimize technical artifacts from low-quality cells and potential doublets, cells meeting the following criteria were filtered out: (1) < 1000 or > 25000 unique molecular identifiers (UMIs, representing unique mRNA transcripts); (2) < 500 or > 5000 genes; or (3) > 10% UMIs derived from the mitochondrial genes. The scanpy’s external module Scrublet (Wolock et al., 2019) was called using the sc.external.pp.scrublet function to identify potential doublets using default parameters. An automatically set threshold labelled 299 cells with a doubletScore > 0.25 as “predicted_doublets” and were filtered out. After quality control, a total of 585860 cells remained. The violin distribution and scatter plot for computed quality measures including gene counts per cell, UMI counts per cell and mitochondrial gene percentage are shown in Supplementary Fig 1. The gene expression matrix were normalized by library size to 10,000 reads per cell by sc.pp.normalize_total function, so that all cells were comparable in UMI counts. Next, the normalized counts were natural log transformed (X = log(X + 1)) by sc.pp.log1p function. The log transformed expression values were used for downstream analysis.

### Batch effect correction and dataset integration

Gene features with high cell-to-cell variation in the data were prioritized using the sc.pp.highly_variable_genes function (supplemental Fig 1). Briefly, the informative highly-variable genes (HVGs) were selected within each sample separately and merged to select the consensus set of 1,500 top-HVGs. All ribosomal and mitochondrial genes were removed from HVGs as described (Cell 2021 Zhangzemin). The HVGs subset matrix extracted from the full expression matrix was used for downstream integration steps. Then each gene was scaled to unit variance and zero mean and clipped when values exceeded 10.

Integration of different datasets was conducted in the order of dimension reduction by principal component analysis (PCA), batch effect correction using Harmony algorithm (Korunsky et al. (2019), Fast, sensitive and accurate integration of single-cell data with Harmony) and unsupervised clustering using Louvain algorithm (Traag et al., 2019). Specifically, the main axes of variation was identified using the sc.tl.pca function with parameter svd_solver=’arpack’. Dimensionality of the datasets was reduced to 50 PCA components and fed into sc.external.pp.harmony_integrate function implemented in the python package harmonypy. The parameter theta was set as 2.5 for sample for technical covariate correction. Nearest neighbor graph of cells was built using the sc.pp.neighbors function with batch-corrected matrix.

### Cell clustering and annotations

Unsupervised clustering of cells was then computed by sc.tl.louvain at different resolutions using the neighborhood relations of cells. Cluster-specific signature genes were identified using the sc.tl.rank_genes_groups function. Cluster annotation was done manually by matching canonical cell marker genes with Cluster-specific signature genes.

Clustering analysis of cell types consisted of two rounds. The first round (Louvain resolution = 1.2) was performed on all cells and identified 10 major cell types (**Fig 1, S1**). The second round (with Louvain resolution ranging from 0.3 to 1.8) was performed on CD4+/CD8+ T cells, B cells, monocyte and DC cells separately to subdivide each cell type into sub-clusters. These sub-clusters represented distinct immune cell lineages within each major cell type. Each sub-cluster was manually analyzed by domain experts and considered as distinctive enough when they had at least one highly expressed signature gene compared to other cells. The complete list of canonical marker genes and cluster-specific highly expressed signatures are provided in Fig 3, 6, 8, S3, S6 and S11.

### Cell state score of cell subtypes

After cluster annotation were completed, several gene sets from important immune processes were used to compare overall activation level or physiological activity of cell clusters. Gene sets related to cytokine storm and immune exhaustion were collected from previous literature (Nature Immun 2020, Cell 2021 zhangzemin) and gene sets about Response To Interferon Alpha (GO:0035455), Response To Interferon Beta (GO:0035456), Acute Inflammatory Response (GO:0002526), Apoptotic Signaling Pathway (GO:0097190), Leukocyte Migration (GO:0050900) were collected from the MsigDB database. Cell state scores were calculated using the sc tl.score_genes function available in Scanpy.

The cell scores of the cell were defined as the average expression of the genes from the predefined gene set with respect to reference genes. Comparison of the cell state score of one condition versus another condition was statistically assessed using the Mann-Whitney rank test (two-tail, p-value < 0.01, adjusted using the Benjamini–Hochberg method).

### TCR and BCR V(D)J immune repertoire sequencing and analysis

From one aliquot of the gene expression 5 libraries, full-length TCR/BCR V(D)J segments were enriched from transcriptome cDNA via PCR amplification using the Chromium Single-Cell V(D)J Enrichment kit according to the manufacturer’s protocol. Similar to the gene expression pipeline, immune repertoire preprocessing was performed using Cell Ranger (v.6.0.0) *vdj* command with human vdj reference vGRCh38-alts-ensembl-5.0.0. This pipeline includes demultiplexing by index and barcodes, TCR/BCR V(D)J sequence discovery and TCR/BCR clonotype assignment to each cell. V(D)J immune repertoire was analyzed by the python-toolkit, Scirpy. In brief, the productive chains of each cell were identified and connected with the cell’s barcode information. Each unique TRA(s)-TRB(s) pair was defined as a TCR clonotype and each unique IGH(s)-IGK/IGL(s) pair was defined as a BCR clonotype. If one clonotype was present in at least two cells, cells harboring this clonotype were considered to be clonal and the number of cells with such pairs indicated the degree of clonality of the clonotype. Only cells with at least one productive clonotype was used in the following analysis. The TCR/BCR downstream analysis were similar for the most part. TCR/BCR data table of cells loaded by Scirpy was matched together with gene expression profiles already prepared by Scanpy in the AnnData data structure. Clonotypes were then clustered based on the similarity of their CDR3 amino acid sequences. TCR/BCR diversity metric, containing clonotype frequency and clonotype composition, was obtained using scirpy.pl.alpha_diversity function based on ‘normalized_shannon_entropy’.

## Statistics

The statistical analysis, visualization and method details described in this study were performed in python and R and are provided with the results of the main text, in the figure legends or in the above Methods sections. In all figures with significance marks, the following convention for symbols indicating statistical significance were used:

ns: p > 0.05

*: p <= 0.05

**: p <= 0.01

***: p <= 0.001

****: p <= 0.0001

## Code availability

Experimental protocols and the data analysis pipeline used in our work follow the 10X Genomics and Seurat official websites. The analysis steps, functions and parameters used are described in detail in the Methods section. Custom scripts for analyzing data are available upon reasonable request. The software and algorithms used in this report are presented in Table S8.

## Funding

This work was supported by grants from State Key Laboratory of Infectious Disease Prevention and Control (2020SKLID303), Beijing Natural Science Foundation (5214023, M21005), Public service development and reform pilot project of Beijing Medical Research Institute (BMR2019-11), National natural science foundation of China (81970900) and Beijing Social Science Foundation Project (19GLB033).

### Ethical approval

This study was approved by the Ethics Committee of the Sanya People’s Hospital (SYPH-2021-26).

### Transparency declaration

The lead author and guarantor affirm that the manuscript is an honest, accurate, and transparent account of the study being reported; that no important aspects of the study have been omitted; and that any discrepancies from the study as planned and registered have been explained.

## Data Availability

Custom scripts for analyzing data are available upon reasonable request.

## Acknowledgments

We thank all the participants. We gratefully acknowledge the participation of Analytical BioSciences Co., Ltd. (Beijing) for the support of construction of single cell sequencing Library, and thanks Miss. RanRan Gao (Analytical BioSciences), JuanJuan Yu (Analytical BioSciences) and Mr. Feng Zhu (Analytical BioSciences) for her contribution.

