## Supplemental Files for "Single-cell transcriptomic atlas of individuals receiving inactivated COVID-19 vaccines reveals distinct immunological responses between vaccine and natural SARS-CoV-2 infection"

**Title Page**

### Figure Legends

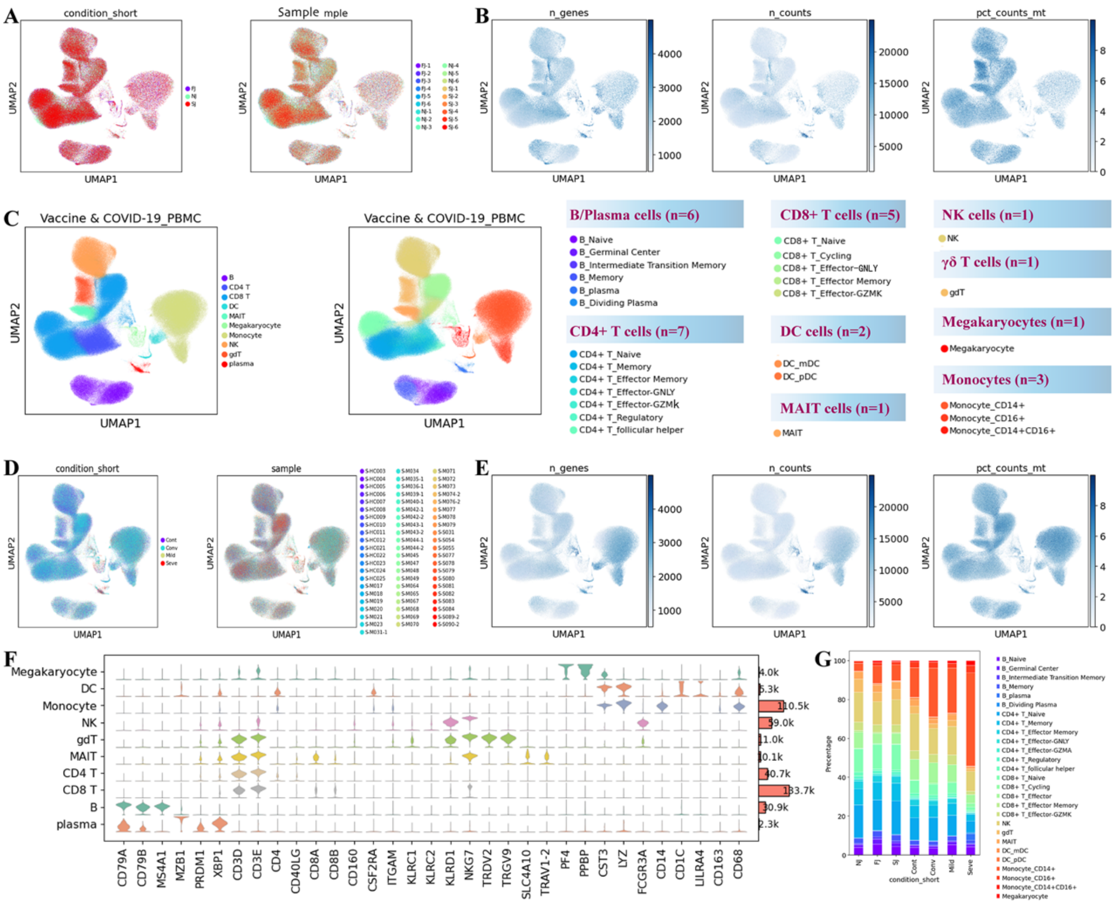


**Fig S1**. **Detailed data output and visualization of single-cell transcriptional profiling of PBMCs from vaccine recipients and COVID-19 infection data sets.**

**A**. The UMAP projection of the vaccine cohort highlighting the three experimental groups (NJ, FJ and SJ) on the left panel and 18 samples on the right panel with different colors.

**B**. The UMAP projection of scRNAseq quality check metrics from the vaccine cohort, including total genes detected for each cell, total transcript detected and mitochondrial gene percentage. The color was linearly correlated with metric values in each plot.

**C**. Cell populations identified and 2-D visualization. The UMAP projection of 590k single cell transcriptomes from both our vaccine cohort and Ren et al. 2021 COVID-19 infection samples, showing the presence of 10 major clusters and 27 smaller clusters with their respective colors.

**D**. The UMAP projection of the COVID-19 cohort highlights the four experimental groups (Cont, Conv, Mild and Seve) on the left panel and 64 samples on the right panel with different colors.

**E**. Similar to B, the UMAP projection of scRNAseq quality check metrics from the COVID-19 infection cohort.

**F**. Expression distribution of cell identity specific RNA markers of the COVID-19 infection cohort. The rows represent 10 cell clusters labeled with different colors and the columns represent log transformed gene expression of the RNAs. The distribution of a gene in a cluster is shown as one small violin plot.

**G.** Composition of cells shown in a percentage stacked barplot. The columns represent seven conditions (NJ, FJ, SJ, Cont, Conv, Mild and Seve) and the colors represent identified cell subtypes.

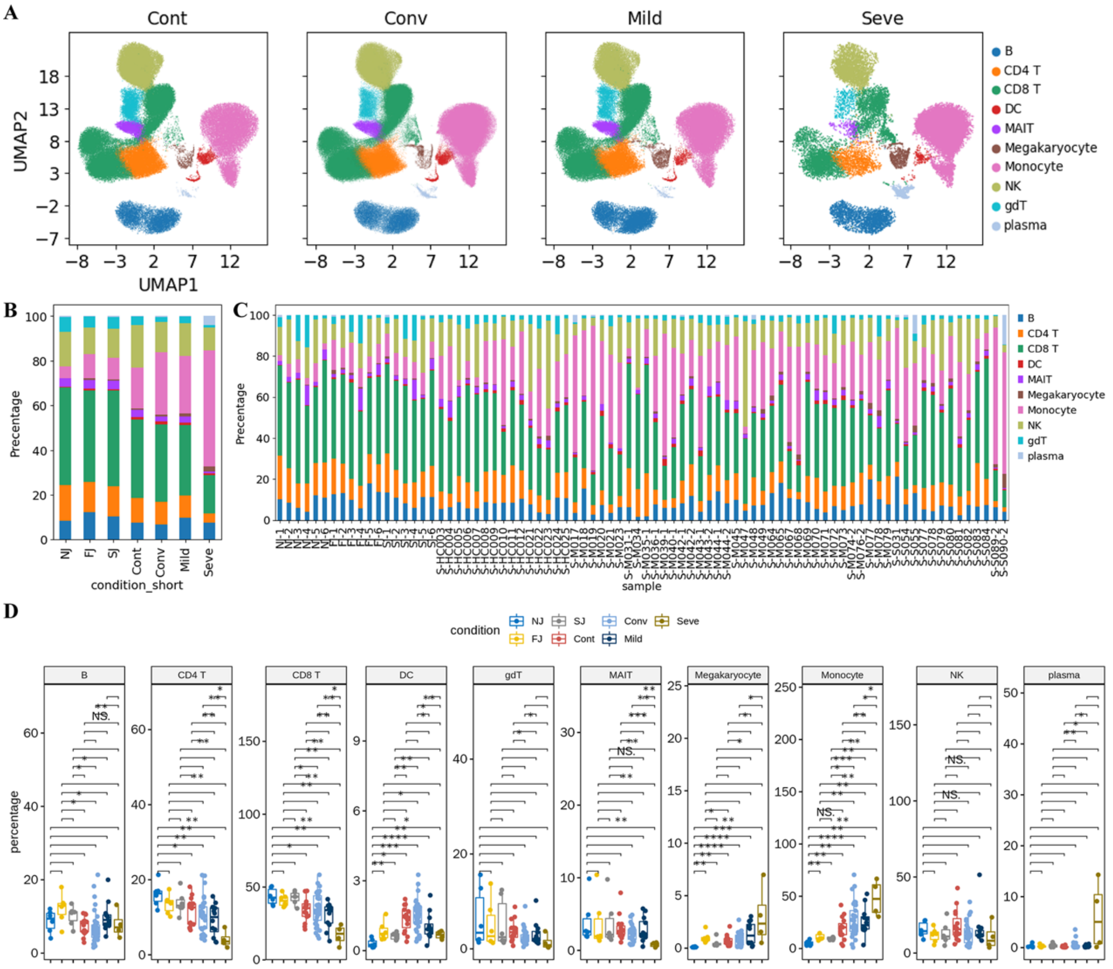


**Fig S2. Additional details about comparison of cell composition across sample groups.**

**A.** UMAP projection of the single cells from Cont, Conv, Mild, and Seve conditions. Each dot corresponds to a single cell, colored by its major cell type.

**B.** Average proportion of each major cell type derived from both vaccine and COVID-19 infection cohorts.

**C.** Proportion of each major cell type derived from each individual vaccine and COVID-19 infection sample.

**D.** The box plot shows cell compositions of both vaccine and COVID-19 infection cohort at a single sample level. All pairwise differences with $P < 0.05$ using two-sided unpaired Mann–Whitney U-test are marked to show significance levels.

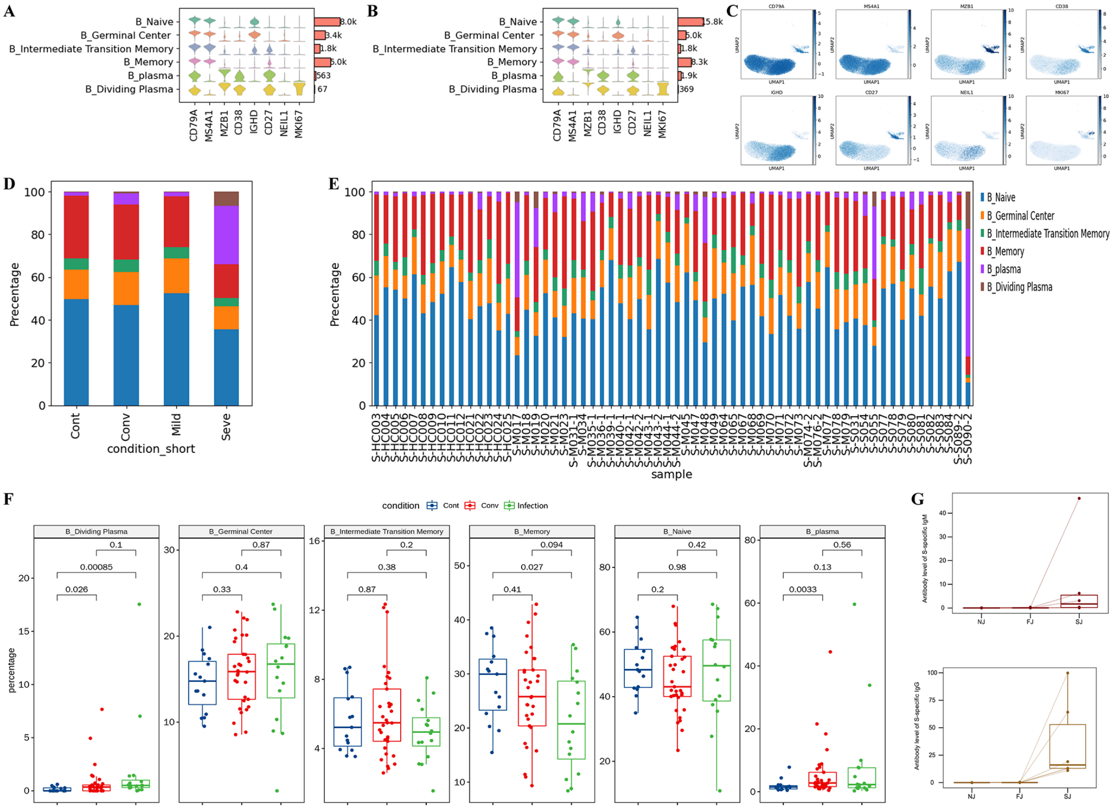

**Fig S3**, **Additional details about the cell type composition of B cells in individuals from vaccination and infection conditions**

**A.** Expression distribution of cell identity specific RNA markers in B cells from the vaccine cohort. The rows represent 5 types of B cells labeled with different colors and the columns represent log transformed gene expression of the RNAs. The distribution of a gene in a cluster is shown as one small violin plot.

**B.** Expression distribution of cell identity specific RNA markers in B cells from the COVID-19 infection cohort.

**C.** Expression levels of canonical B cell RNA markers were used to identify and label major cell clusters on the UMAP plot. Cells are colored according to the log transformed and normalized expression levels of eight genes. Cells are from the COVID-19 infection cohort.

**D.** Average proportion of each B cell subtype derived from COVID-19 infection cohort.

**E.** Proportion of each B cell type derived from each individual vaccine and COVID-19 infection sample.

**F.** The box plot shows B cell compositions of Cont, Conv, Infection (Mild and Seve) conditions at a single sample level.

**G.** Serum antibody levels of IgM and IgG in NJ, FJ and SJ conditions from six participants.

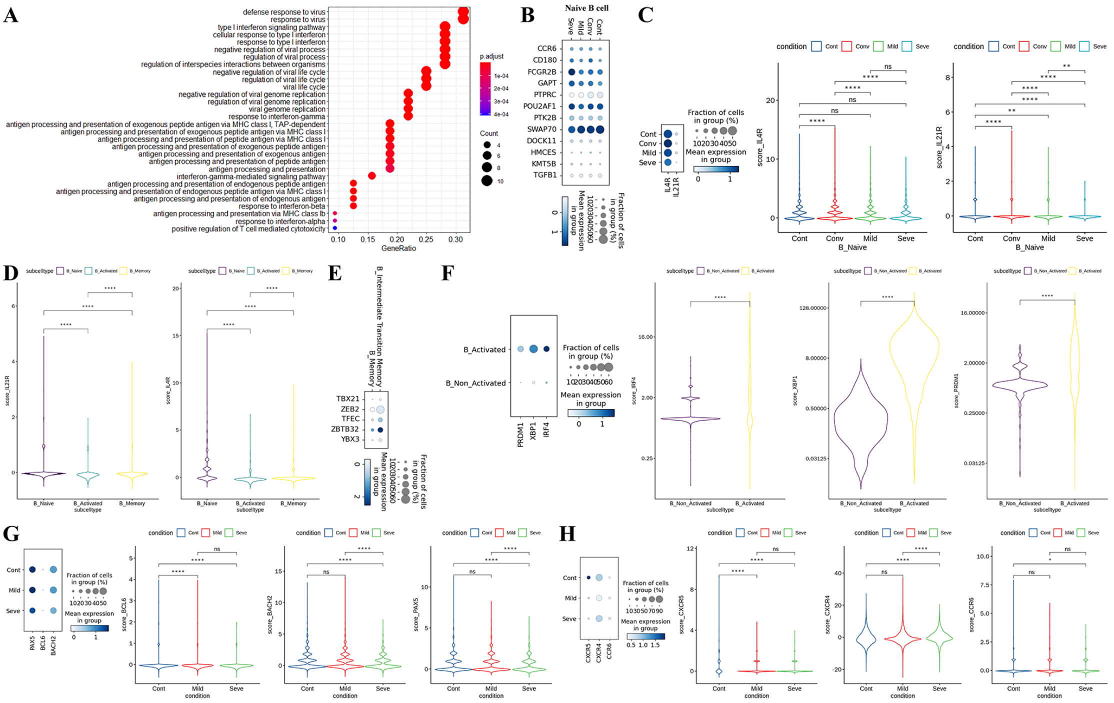


**Fig S4. Characterization of gene expression differences in B cells in individuals across COVID-19 infection conditions.**

**A.** GO enrichment analysis of DEGs identified by comparing control and infected conditions. DEGs refer to genes with Benjamini–Hochberg adjusted P value (two-sided unpaired Mann–Whitney U-test) ≤0.01 and average log2 fold change ≥1 in both Mild/Cont and Seve/Cont comparisons.

**B.** Dot plots of the gene expression level of naïve B cells in Cont, Conv, Mild and Seve conditions. Columns represent conditions; rows represent genes. Dots are colored by mean expression levels in each condition.

**C.** Violin plots show the expression levels of *IL4R* and *IL21R* in naïve B cells from Cont, Conv, Mild and Seve conditions.

**D.** Violin plots show the expression levels of *IL4R* and *IL21R* in naïve B, cells activated B cells and memory B cells.

**E.** Dot plots of gene expression level of memory B cells and intermediate transition memory B cells in the COVID-19 infection cohort. Rows represent genes (*TBX21*, *ZEB2*, *TFEC*, *ZBTB32*, *YBX3*); columns represent B cell subtypes. Dots are colored by mean expression levels in each group.

**F.** *PRDM1*, *XBP1* and *IRF4* gene expression level of activated B cells and other B cells in the COVID-19 infection cohort. Dot plots (Left) and violin plots (Right) are used for visualization.

**G.** *PAX5*, *BCL6* and *BACH2* gene expression level of B cells in Cont, Conv, Mild and Seve conditions. Dot plots (Left) and violin plots (Right) are used for visualization.

**H.** *CXCR5*, *CXCR4* and *CCR6* gene expression level of B cells in Cont, Conv, Mild and Seve conditions. Dot plots (Left) and violin plots (Right) are used for visualization.

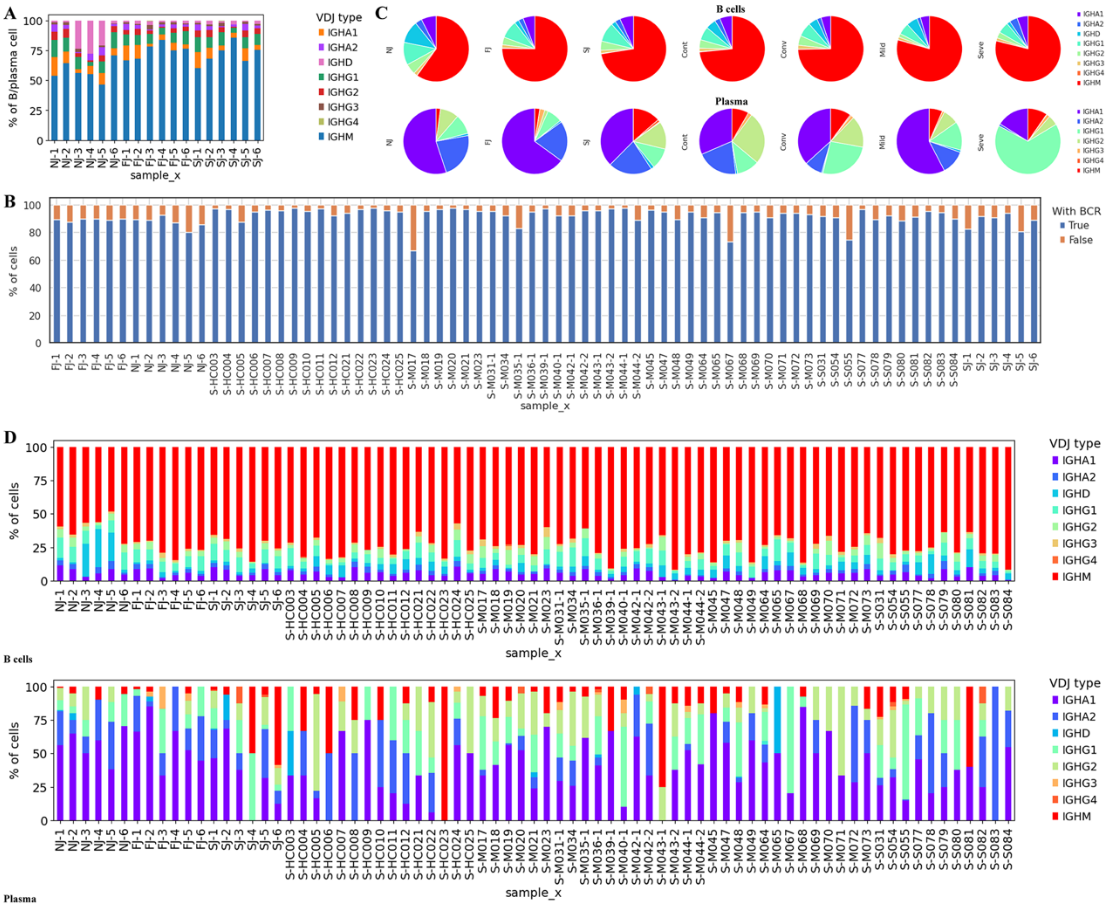


**Fig S5. Additional information about BCR clones analysis.**

**A**, Stacked bar plots show the percentages IGHA, IGHD, IGHG and IGHM in both B cells and plasma cells from each sample.

**B**, Stacked bar plots show the BCR detection rate in each sample.

**C**, Pie graphs showing the distribution of IGHA, IGHD, IGHG and IGHM in B cells and plasma cells from each condition.

**D**, Stacked bar plots showing the percentages IGHA, IGHD, IGHG and IGHM in B cells and plasma cells at the sample level.

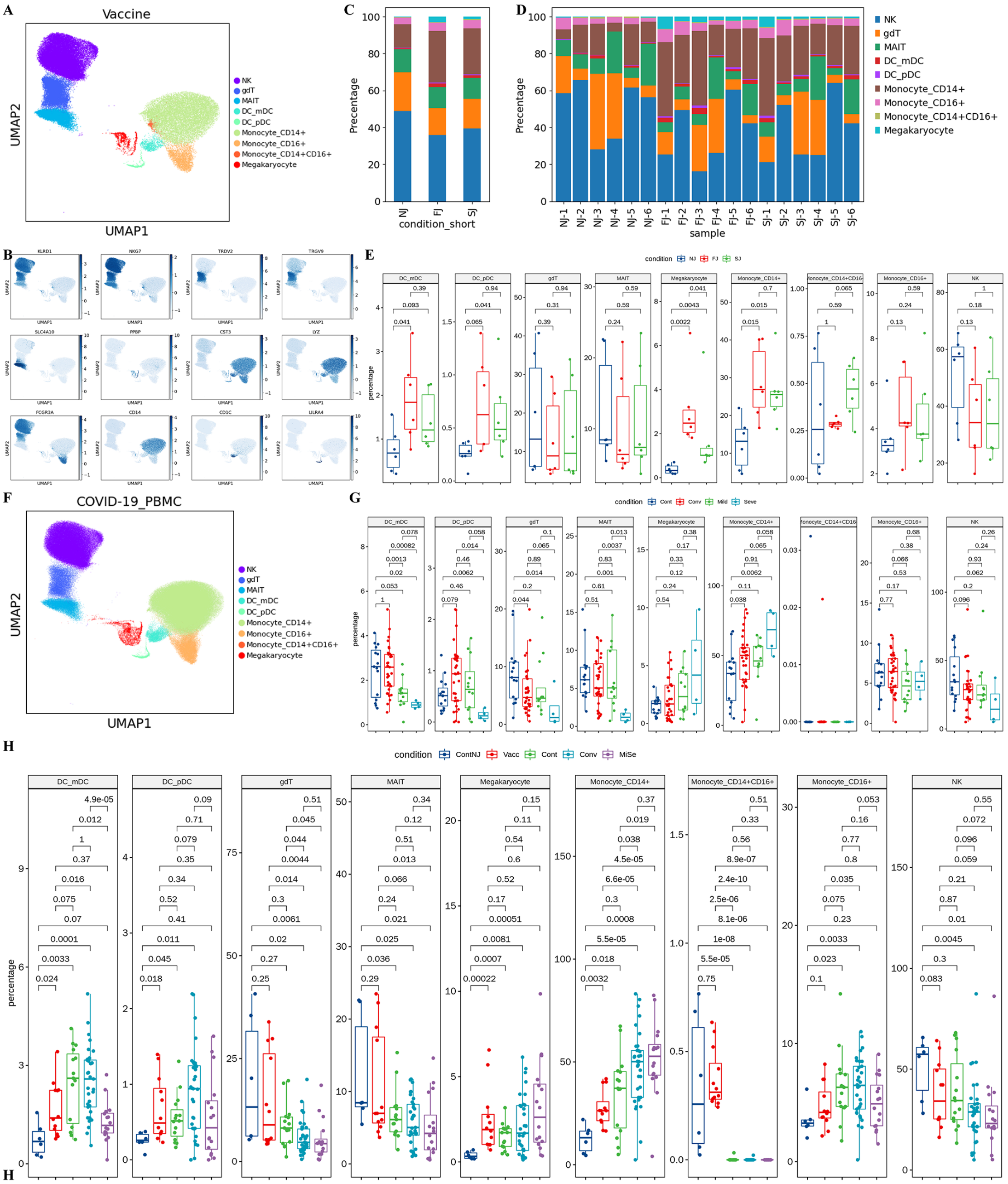


**Fig S6. Additional information about characterization of innate cell composition differences in individuals across vaccine and infection conditions.**

**A**, Expression distribution of cell identity specific RNA markers of vaccine cohort samples. The rows represent 9 innate cell clusters labeled with different colors and the columns represent log transformed gene expression of the RNAs. The distribution of a gene in a cluster is shown as one small violin plot.

**B**, Expression levels of canonical single cell RNA markers were used to identify and label cell clusters on the UMAP plot. Cells are colored according to log transformed and normalized expression levels of twelve genes, KLRD1, NKG7, TRDV2, etc.

**C**, Expression distribution of cell identity specific RNA markers of COVID-19 cohort samples. The rows represent 9 innate cell clusters labeled with different colors and the columns represent log transformed gene expression of the RNAs. The distribution of a gene in a cluster is shown as one small violin plot.

**D**, Average proportion of each innate cell subtype derived from Cont, Conv, Mild and Seve groups.

**E**, Percentage of each innate cell subtype derived from Cont, Conv, Mild and Seve individual samples.

**F**, Average proportion of each innate cell subtype derived from NJ, FJ, SJ, Cont, Conv, Mild and Seve groups.

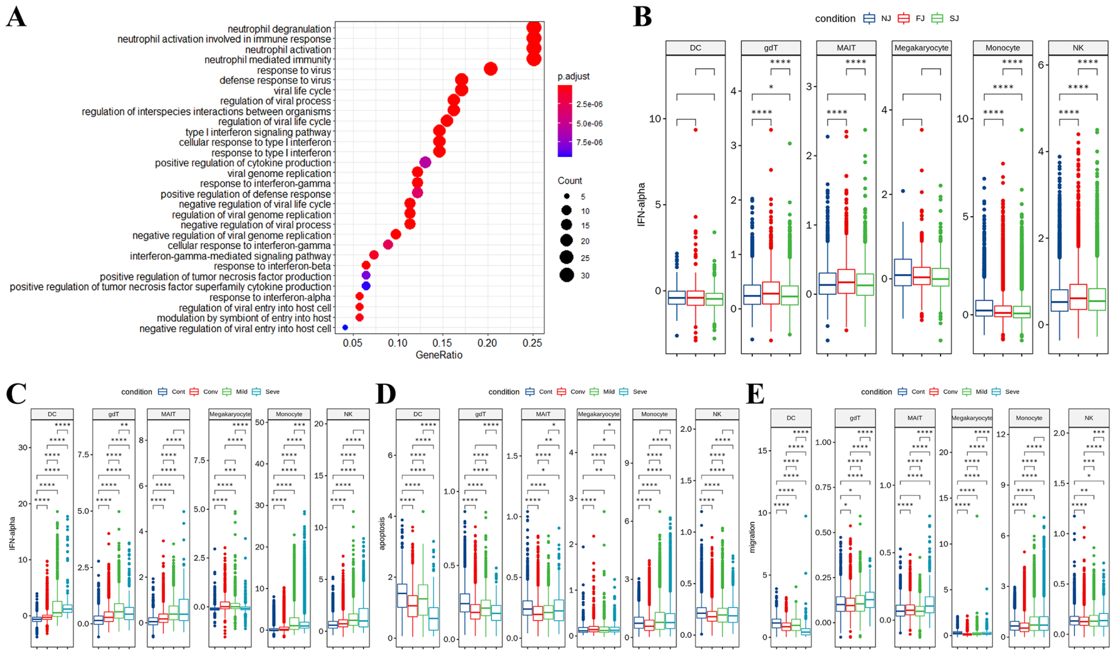


**Fig S7, Characterization of gene expression differences in innate immune cells from vaccine and COVID-19 cohort samples.**

**A**, GO enrichment analysis of DEGs identified by comparing the COVID-19 infection and healthy control conditions. DEGs refer to genes with Benjamini–Hochberg adjusted P value (two-sided unpaired Mann–Whitney U-test) ≤0.01 and average log2 fold change ≥1 in both Mild/Cont and Seve/Cont comparisons.

**B**, Expression activity of IFN-alpha pathways in NJ, FJ and SJ conditions shown as box plots. Boxes are colored by vaccine conditions.

**C**, **D**, **E**, Expression activity of IFN-alpha, apoptosis and migration pathways in Cont, Conv, Mild and Seve conditions shown as box plots.

All pairwise differences with $P < 0.05$ using two-sided unpaired Mann–Whitney U-test are marked to show significance levels.

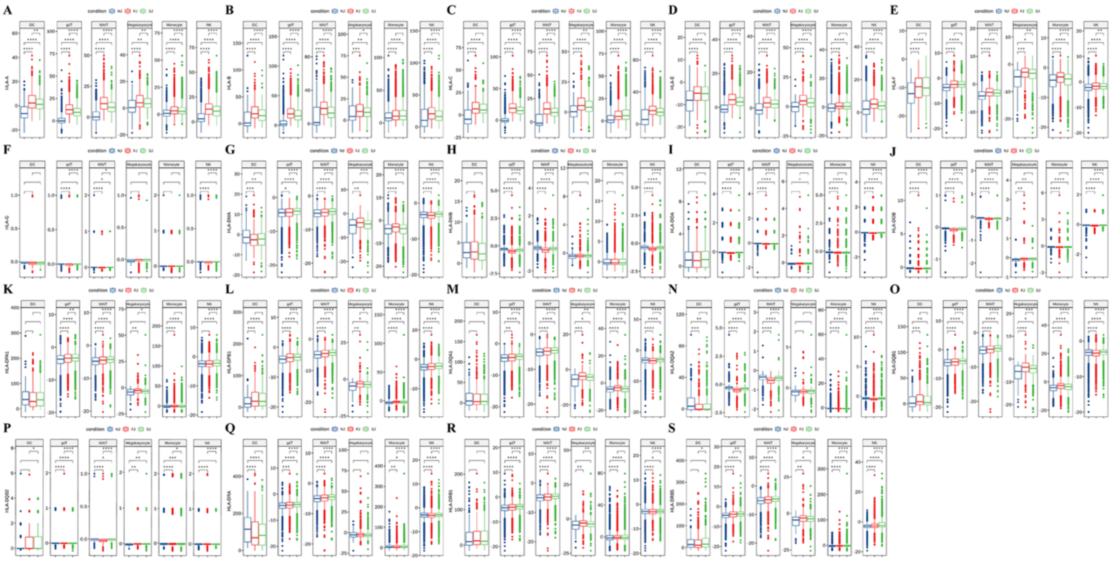

**Fig S8. Expression of HLA-I and HLA-II genes in innate cells from NJ, FJ, and SJ conditions at the sample level.**

A-S are box plots for each HLA gene as indicated in the y axis label.

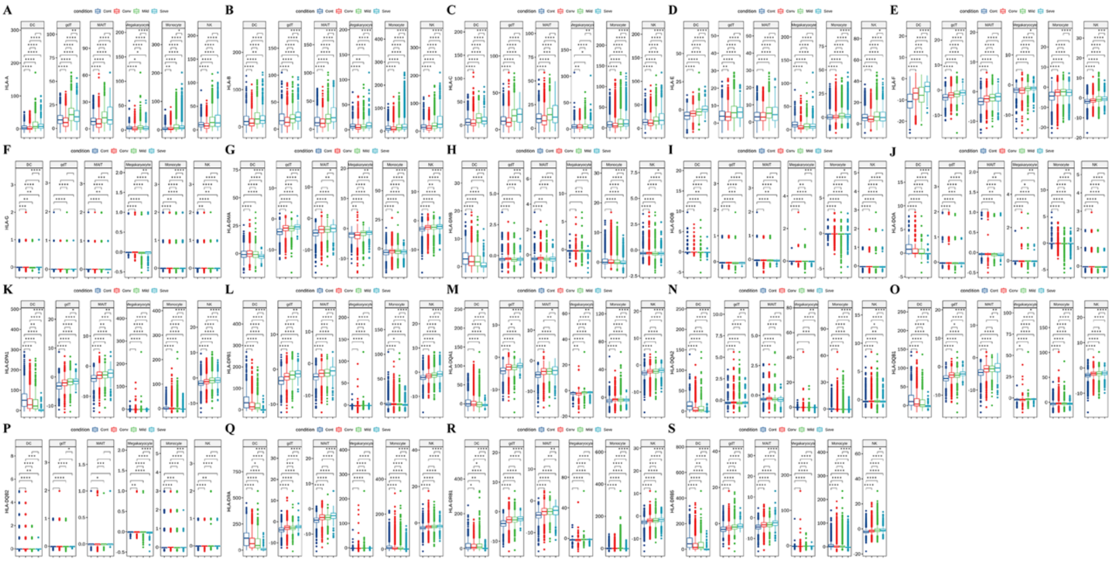

**Fig S9. Expression of HLA-I and HLA-II genes in innate cells from Cont, Conv, Mild and Seve conditions at the sample level.**

A-S are box plots for each HLA gene as indicated in the y axis label.

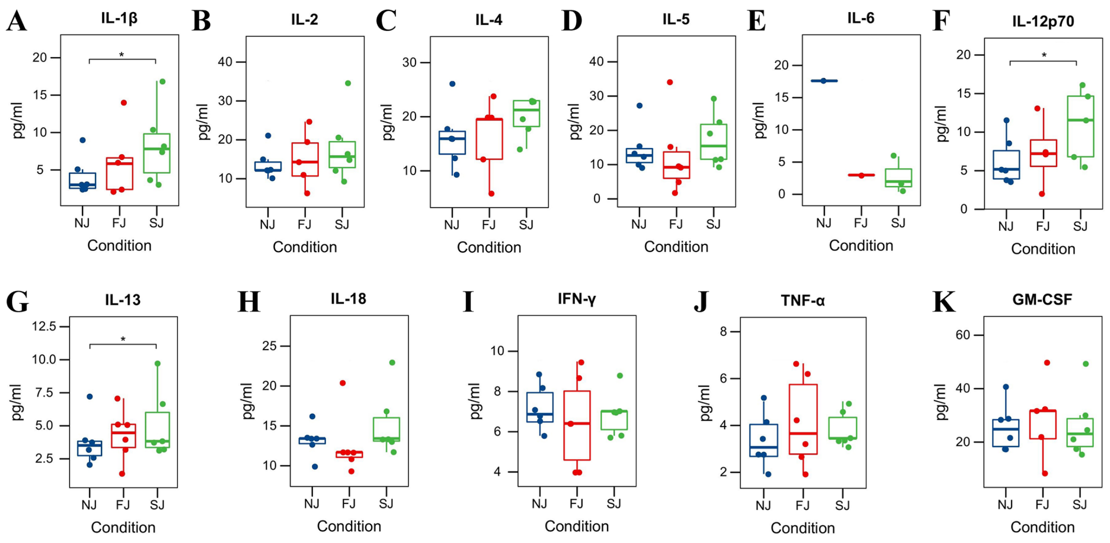


**Fig S10. Cytokine protein levels in the sera of vaccine cohort samples.**

Box plots A-K show the protein levels of 11 cytokines, including IL-1beta, IL-2, etc.

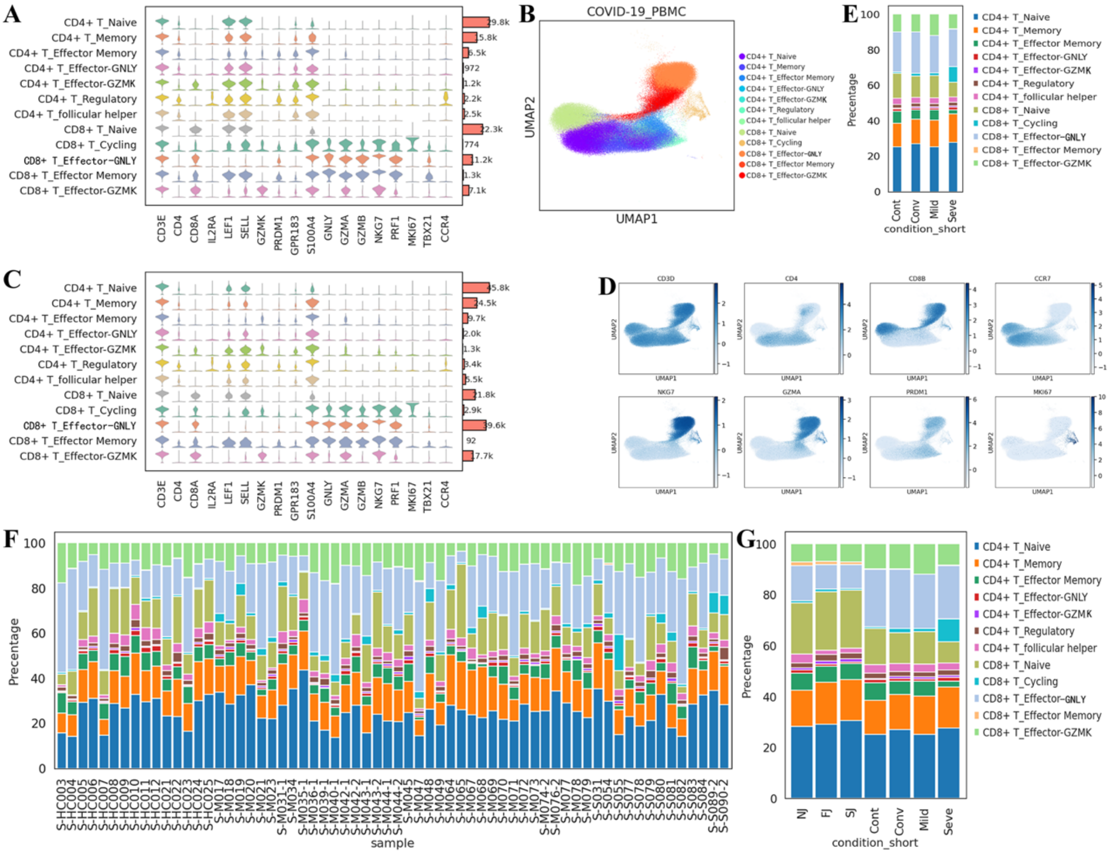


**Fig S11. Additional details about the cell type composition of T cells in individuals from vaccine and COVID-19 infection conditions.**

**A**, Expression distribution of cell identity specific RNA markers in T cells of the vaccine cohort. The rows represent subtypes of T cells labeled with different colors and the columns represent log transformed gene expression of the RNAs. The distribution of a gene in a cluster is shown as one small violin plot.

**B**, UMAP projection of T cells from the COVID-19 infection cohort. The T cells are colored by cell subtypes.

**C**, Expression distribution of cell identity specific RNA markers in T cells of the COVID-19 infection cohort.

**D**, Expression levels of canonical T cell RNA markers were used to identify and label major cell clusters on the UMAP plot. Cells are colored according to the log transformed and normalized expression levels of eight genes. Cells are from COVID-19 infection cohort.

**E**, Average proportion of each T cell subtype derived from COVID-19 infection cohort.

**F**, Stacked bar plots show the proportion of each T cell subtype derived from each individual vaccine and COVID-19 infection sample.

**G**, Stacked bar plot shows the average composition of T cell subtypes of Cont, Conv, Mild and Seve conditions at the group level.

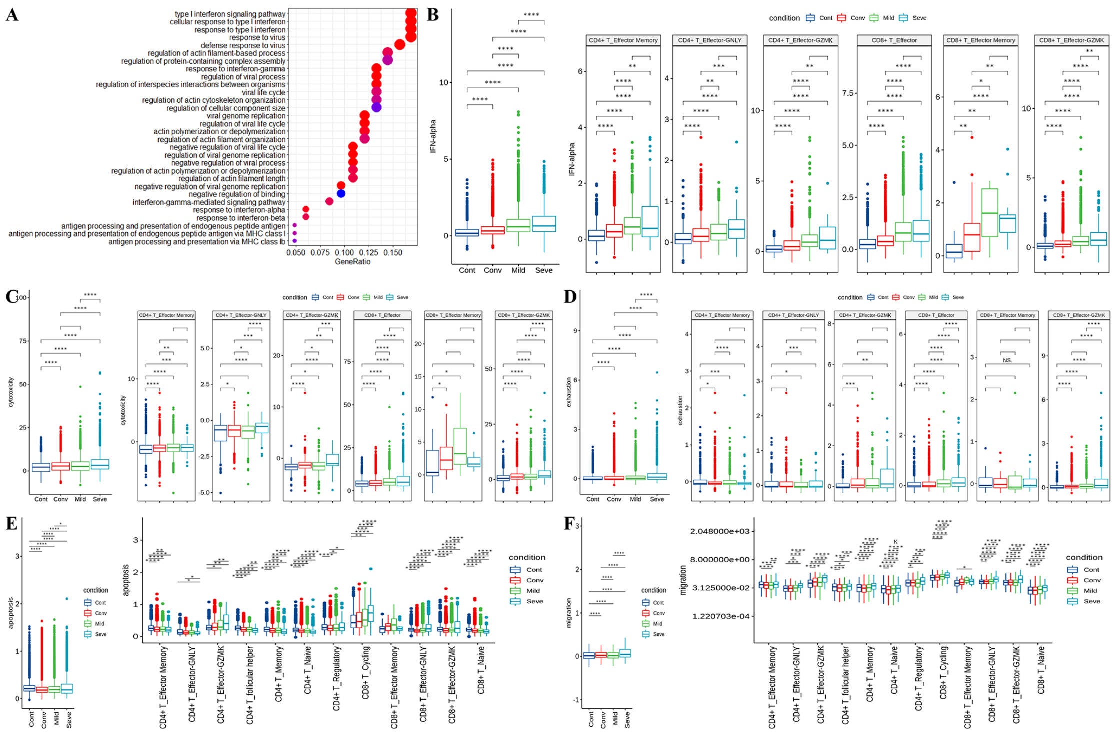


**Fig S12. Characterization of gene expression differences in activated T cells from COVID-19 cohort samples.**

**A**, GO enrichment analysis of DEGs identified by comparing the before vaccine or after vaccine conditions. DEGs refer to genes with Benjamini–Hochberg adjusted P value (two-sided unpaired Mann–Whitney U-test) ≤0.01 and average log2 fold change ≥1 in both Mild/Cont and Seve/Cont comparisons.

**B**, Expression activity of IFN-alpha pathways in activated T cells (Left) and subtypes (right) of Cont, Conv, Mild and Sere conditions shown as box plots and are colored by sample conditions.

**C**, Expression activity of cytotoxicity pathways in activated T cells (Left) and subtypes (right) of Cont, Conv, Mild and Sere conditions shown as box plots and are colored by sample conditions.

**D**, Expression activity of exhaustion genes in activated T cells (Left) and subtypes (right) of Cont, Conv, Mild and Sere conditions shown as box plots and are colored by sample conditions.

**E**, Expression activity of apoptosis pathways in activated T cells (Left) and subtypes (right) of Cont, Conv, Mild and Sere conditions shown as box plots and are colored by sample conditions.

**F**, Expression activity of migration pathways in activated T cells (Left) and subtypes (right) of Cont, Conv, Mild and Sere conditions shown as box plots and are colored by sample conditions.

All pairwise differences with $P < 0.05$ using two-sided unpaired Mann–Whitney U-test are marked to show significance levels.

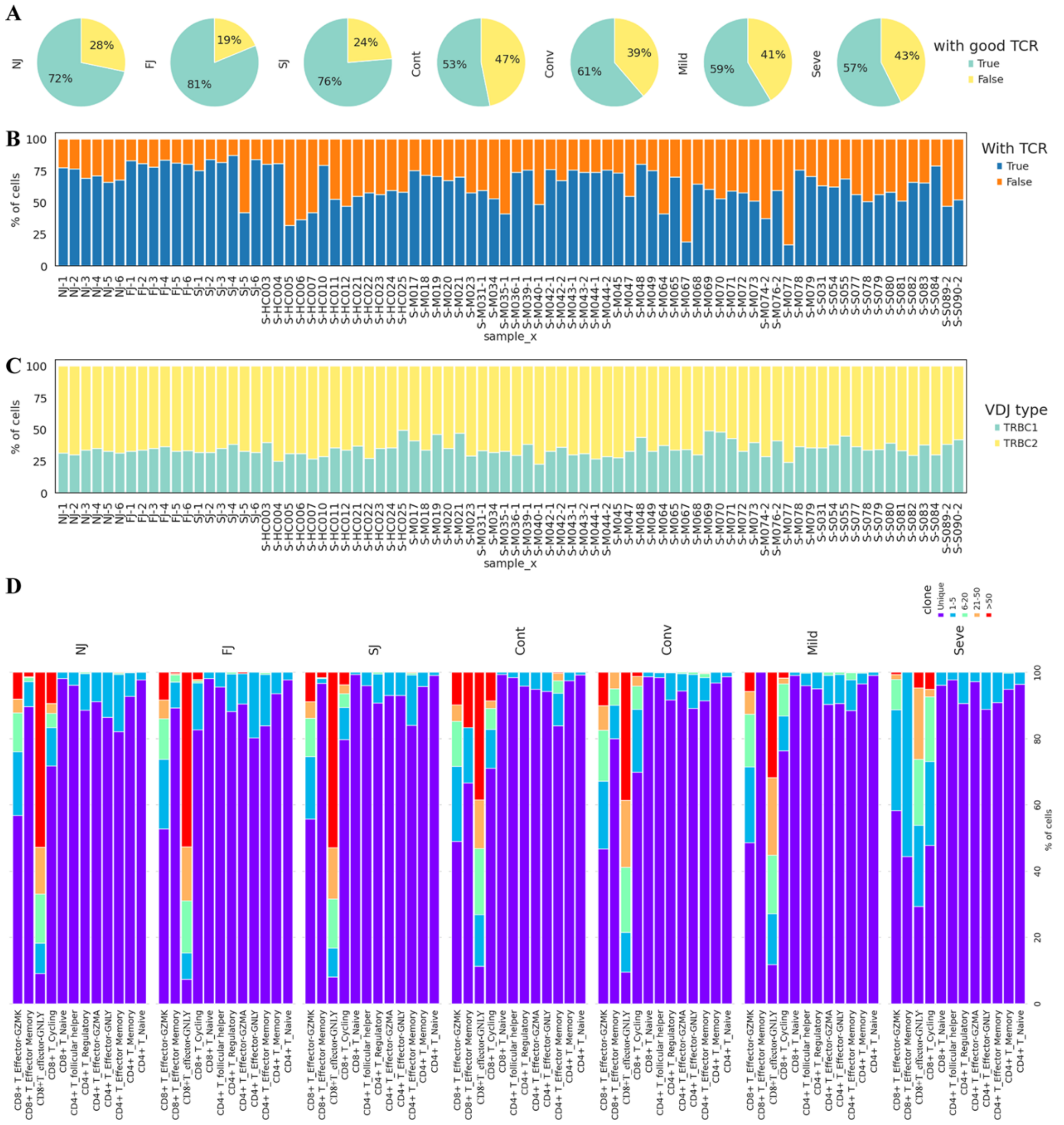


**Fig S13. Additional figures for TCR analysis.**

**A**, Pie graphs show the TCR detection rate of T cells for each condition.

**B**, Stacked bar plots show the TCR detection rate in T cells for each conditions at the single sample level.

**C**, Stacked bar plots show the TCR VDJ type composition in T cells for each condition at the single sample level.

**D**, Stacked bar plots show the TCR clone expansion in cell subtype of T cells in seven conditions.

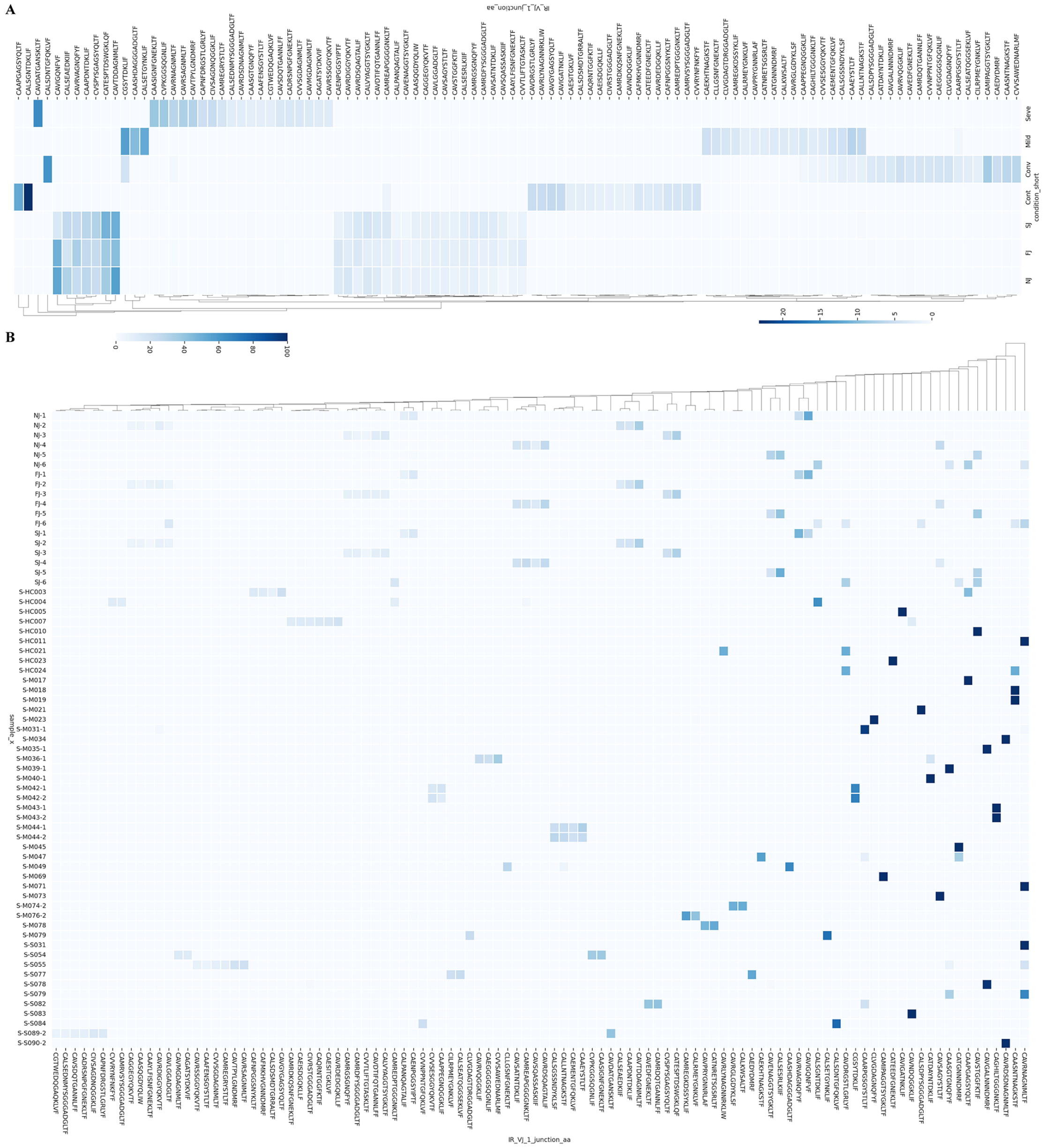

**Fig S14. TCR VDJ junction amino acids sequences.**

**A**, Usage heatmap of each amino acid sequence shown as the condition normalized percentage value. The amino acid sequences are hierarchically clustered by usage similarity.

**B**, Usage heatmap of each amino acid sequence shown as the sample normalized percentage value. The amino acid sequences are hierarchically clustered by usage similarity.
